# Decomposing growth in a national HL7 CDA clinical document repository

**DOI:** 10.64898/2026.05.24.26353991

**Authors:** Harry-Anton Talvik, Sven Laur, Jaak Vilo, Sulev Reisberg

**Affiliations:** Institute of Computer Science, University of Tartu, 51009 Tartu, Estonia; STACC, 51009 Tartu, Estonia

## Abstract

Longitudinal evaluations of national electronic health record repositories often track document counts alone, obscuring changes in content size, structure and standards implementation. We decomposed growth in the Estonian Health Information System across document counts, per-document size, section-level structure and version uptake in a 10% random population sample of 4.97 million HL7 Clinical Document Architecture Release 2 documents from 147,819 patients, spanning 2012–2019 and four prespecified document types. Growth patterns differed by document type. Inpatient summaries increased 48.5% in total content volume despite a 2.4% decline in document counts. Section presence and within-section content were highly skewed; 44.6% of 892 data locations carried one fixed value. Code-system diversity increased from 45 to 79, and version uptake took years: inpatient summaries reached 80% organisational uptake after a median 44 months (95% CI 11–78). This decomposition can guide extraction pipelines, secondary use and standards governance in CDA- and FHIR-based repositories.

## Introduction

National electronic health record (EHR) systems generate clinical documents at an increasing scale, and recent longitudinal studies have begun to quantify specific dimensions of this growth. At the document level, median outpatient progress note length at a large US academic medical centre increased by 60% between 2009 and 2018 (2,704,800 notes across 46 specialties), a compound annual growth rate (CAGR) of approximately 4.8% [1]. At the encounter level, the median number of prior notes available to emergency physicians at another large US health system rose from 5 in 2006 to 359 in 2022 (730,968 encounters), reflecting cumulative note burden rather than new per-year production [2]. In parallel, clinical terminologies such as SNOMED CT, LOINC, and RxNorm have expanded steadily, each adding thousands of concepts per year [3]. Yet each of these studies addresses only a single dimension of growth, typically within a single institution or at the specifications level rather than in production document repositories. No study has examined how longitudinal growth decomposes below the document level in national health information systems, where document structure, coding practices, and implementation standards all evolve at the same time.

We address this gap with a full-extraction analysis of Health Level Seven (HL7) Clinical Document Architecture Release 2 (CDA R2) documents from the Estonian Health Information System (EHIS). Estonia operates a nationwide, mandatory electronic health record platform that has been collecting CDA R2 documents from all healthcare providers since 2009 [4]. The system integrates data from over a thousand healthcare institutions via the national X-Road data exchange infrastructure, and by 2017 contained more than 30 million clinical documents covering the entire resident population [4]. Our analysis uses the RITA MAITT dataset, a 10% random sample of the Estonian population covering 147,819 patients and approximately 5 million documents from 2012 to 2019 [5]. Because this period reflects system maturation rather than current practice, we focus on structural and methodological patterns that generalise beyond this particular period.

The underlying question is practical: how should extract, transform, and load (ETL) pipeline maintainers allocate monitoring and development effort across document sections, value locations in Extensible Markup Language (XML) documents addressed by XML Path Language (XPath) expressions, and schema versions? The same decomposition informs standards governance and data-availability assessment for secondary use. The paper is organised around four results that directly address these questions: the first two concern where within documents to concentrate monitoring effort, and the latter two concern how to plan across schema versions. First, health data growth is driven by several concurrent components rather than a single document-count trend. Second, section presence and field completeness are highly heterogeneous. Third, schema evolution is gradual and continuous rather than neatly segmented into versioned breaking points. Fourth, standard adoption is slow enough that multiple document versions coexist for years across organisations.

## Results

### Dataset and corpus characteristics

The dataset comprised 4,971,271 CDA R2 documents generated between 1 January 2012 and 31 December 2019, representing a 10% random sample of the Estonian population (147,819 patients). Documents were distributed unevenly across the four analysed types, with outpatient discharge summaries accounting for the majority of the corpus (Table 1). Over the study period, the size of the source population remained broadly stable: according to Statistics Estonia [6], the population of Estonia was 1,325,217 on 1 January 2012 and 1,328,889 on 1 January 2020, a net increase of 3,672 persons (0.28%) despite non-monotonic year-to-year variation.

**Table 1.** Dataset overview. CDA R2 documents from a 10% random sample of the Estonian population, 2012–2019.

|  | Documents, *n* (%) | Patients, *n* | Median file size, kB (IQR) |
| --- | --- | --- | --- |
| Outpatient discharge summary | 2,693,575 (54.2%) | 142,781 | 13.8 (10.9–20.4) |
| Inpatient discharge summary | 168,473 (3.4%) | 67,109 | 109.3 (24.1–224.8) |
| Referral | 465,532 (9.4%) | 106,271 | 10.8 (9.3–13.1) |
| Referral response | 1,643,624 (33.1%) | 130,554 | 10.4 (5.9–20.0) |
| **Total** | 4,971,271<sup>†</sup> (100%) | 147,819<sup>‡</sup> | 12.6 (9.6–20.3) |

IQR, interquartile range. Percentages may not sum to 100 due to rounding. <sup>†</sup> Includes 67 documents that could not be classified into any of the four types due to missing or unrecognised type metadata; these are counted in the total but not in any per-type row. <sup>‡</sup> Unique patients across all document types; individuals may appear in multiple types, so per-type counts do not sum to the total.

The full extraction pipeline parsed all documents into individual data elements. Of the initial 4,971,271 documents, 16,392 (0.33%) were excluded due to malformed XML or empty clinical content, affecting 5,241 patients (103 of whom had no remaining valid documents). The resulting corpus comprised 4,954,879 documents from 147,716 patients and varied substantially in document complexity across types (Table 2).

**Table 2.** Extracted corpus summary. Structural characteristics of the fully parsed CDA R2 corpus. Median field counts reflect extracted field instances per document.

|  | Organisations, *n* | Document size, characters |  | Fields per document |  |
| --- | --- | --- | --- | --- | --- |
|  |  | Median | IQR | Median | IQR |
| Outpatient discharge summary | 876 | 4,132 | (3,232–6,148) | 221 | (184–317) |
| Inpatient discharge summary | 48 | 17,419 | (7,726–30,239) | 1,423 | (312–2,621) |
| Referral | 757 | 3,411 | (2,721–4,255) | 172 | (166–195) |
| Referral response | 64 | 2,446 | (1,910–3,550) | 118 | (96–247) |
| **Total** | 957<sup>†</sup> | 3,577 | (2,656–5,365) | 197 | (150–301) |

IQR, interquartile range. <sup>†</sup> Unique organisations; providers may submit multiple document types. Total extracted field instances: 1,816,065,905 across all documents, including structurally present instances with empty values.

Documents were composed of 31 distinct section types (Table 3). Document hierarchy was shallow: nesting depth did not exceed two levels, and only 0.17% of section instances (35,430 of 21.2 million) were nested. Four section types (`PAT_DGN`, `CLIN_DGN`, `KOLO`, `DRUGSCH`) appeared exclusively as second-level children. By contrast, `DGN` (Final clinical diagnosis) was the only type occurring at both levels: it was nested within `DEC` (Decision) in referral response documents (0.20% of all `DGN` occurrences) but appeared at the top level in all other document types. A parser keying only on section code would therefore conflate final clinical diagnoses from discharge summaries and referrals with the diagnostic content embedded in referral-response decisions, even though these carry different clinical meaning.

**Table 3.**
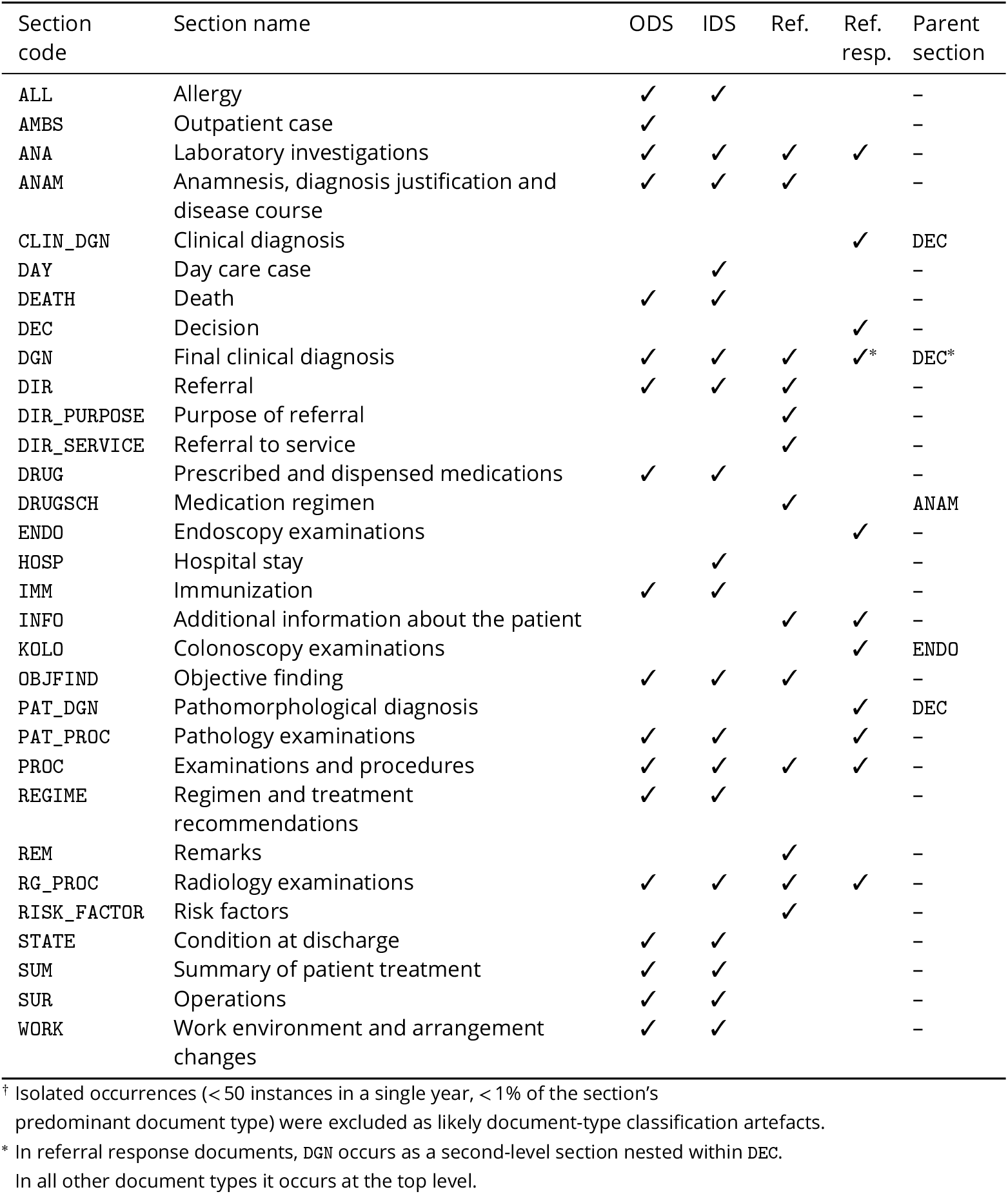
Section types in the dataset. Presence of 31 section types across four document types, 2012–2019.^†^ ODS = Outpatient discharge summary; IDS = Inpatient discharge summary; Ref. = Referral; Ref. resp. = Referral response.

The full inventory of 125 section types defined across all Estonian CDA R2 document specifications, including sections belonging to document types not analysed in this study, is provided in the supplementary information (Supplementary Table S1), together with a section-type introduction timeline (Supplementary Fig. S5).

### Decomposing health data volume growth

Health data volume is the product of three concurrent components: the number of active patients generating records, the number of documents per patient per year, and the amount of data carried by each document. Because these components can evolve independently, aggregate volume trends can conceal distinct underlying dynamics. We therefore examine patient reach, document frequency, and document size in turn, and show that the three combine in markedly different proportions across document types.

#### Patient counts and document frequency

Active patient counts increased over 2012–2019 for referrals, referral responses, and outpatient discharge summaries, whereas inpatient discharge summaries showed a slight decline (Figure 1a). Referrals rose most steeply, with more moderate growth for referral responses and outpatient summaries (Table 4). The steep referral trajectory coincides with the introduction of nine referral subtypes in the national document-type registry on 2014-07-01 [7], which expanded the operational definition of a referral mid-window.

**Table 4.** Average annual percent change (AAPC) from 2012 to 2019, by document type, for the three multiplicative components of health data volume: active patient counts (Figure 1a), mean documents per patient (Figure 1b), and average document size (Figure 2). Confidence intervals are 95% Wald intervals.

| Metric | Document type | AAPC (%) | 95% CI |
| --- | --- | --- | --- |
| Active patients | Outpatient discharge summaries | +9.4 | [5.3, 13.6] |
|  | Inpatient discharge summaries | -1.0 | [-1.5, -0.5] |
|  | Referrals | +99.6 | [61.3, 147.0] |
|  | Referral responses | +14.1 | [11.4, 17.0] |
| Documents per patient | Outpatient discharge summaries | +9.0 | [6.8, 11.1] |
|  | Inpatient discharge summaries | +0.7 | [0.5, 1.0] |
|  | Referrals | +10.1 | [7.5, 12.7] |
|  | Referral responses | +16.0 | [11.4, 20.8] |
| Average document size | Outpatient discharge summaries | +1.3 | [1.0, 1.7] |
|  | Inpatient discharge summaries | +5.5 | [2.6, 8.6] |
|  | Referrals | +3.0 | [1.3, 4.7] |
|  | Referral responses | +5.0 | [2.6, 7.6] |

Note. All 95% confidence intervals exclude 0.

**Figure 1.**
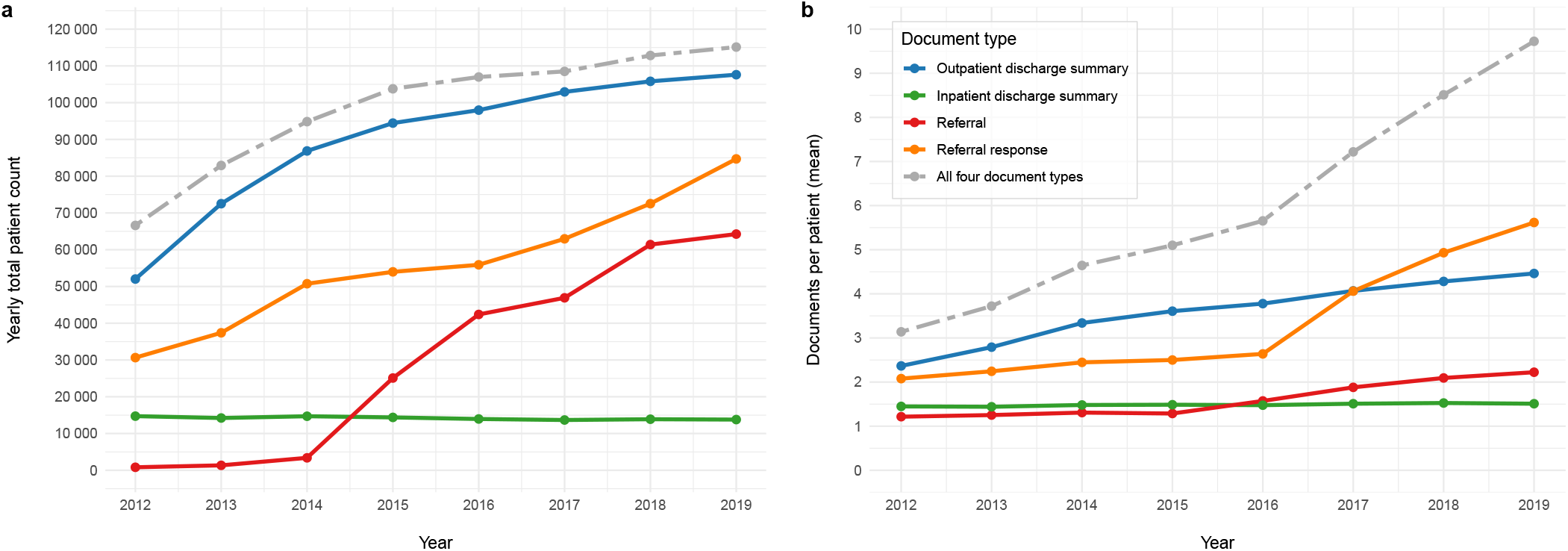
Decomposition of volume growth into patient reach and document frequency. **a**, Active patients per year, stratified by document type. **b**, Documents per patient per year, stratified by document type. Colour and line style encode document type consistently across both panels (legend in **b**).

Document frequency showed a different pattern. Defined as the mean number of documents per patient per year, it increased for all four document types (Figure 1b). The rise was strongest for referral responses and referrals, substantial for outpatient summaries, and minimal for inpatient summaries, which were essentially unchanged (Table 4).

#### Document size

Average document size, measured as the median sum of field-value lengths (in characters) per document, grew for all four types, but at markedly different rates (Figure 2, Table 4). Inpatient discharge summaries grew fastest, followed by referral responses, then referrals. Outpatient discharge summaries grew slowest, at roughly one-quarter the rate of the inpatient and referral-response types.

**Figure 2.**
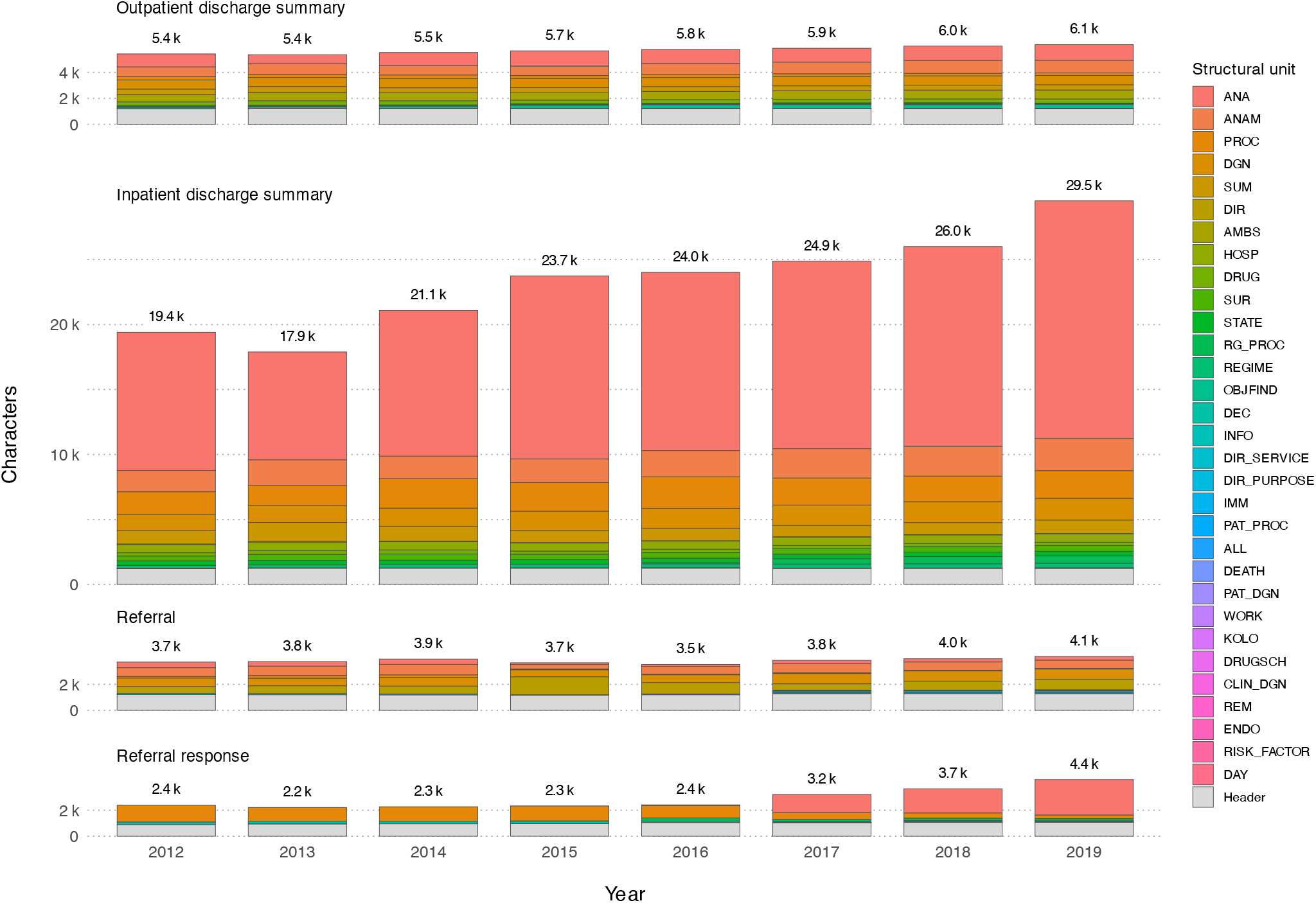
Average document size by year and document type, shown as a stacked decomposition by section type plus header content.

Taken together, the three components combine in contrasting patterns by document type: outpatient summaries grew through patient reach and document frequency, inpatient summaries through document size alone, and referral-related documents through all three simultaneously. The inpatient case is most striking: across 2012–2019, per-document size rose cumulatively by roughly 50%, more than offsetting a −2.4% cumulative decline in document counts to produce a +48.5% cumulative net increase in aggregate inpatient data volume.

### Document structure and section presence

Beyond the document-level volume trends reported above, CDA documents exhibit internal structural heterogeneity that varies across section types, document types, and time. We examine this at two levels: section presence within documents, and location usage within sections.

#### Section-level presence

Figure 3 shows the proportion of inpatient discharge summaries containing each section type, by year. The distribution is highly skewed: of 21 observed section types, only 8 exceed 50% presence in any year. Four sections—`HOSP`, `DGN`, `ANAM`, and `STATE`—appear in more than 99% of documents throughout, while a second tier (`SUM`, `ANA`, `REGIME`, `PROC`) occurs in 50% to 90%. The remaining section types each appear in fewer than 40% of documents.

**Figure 3.**
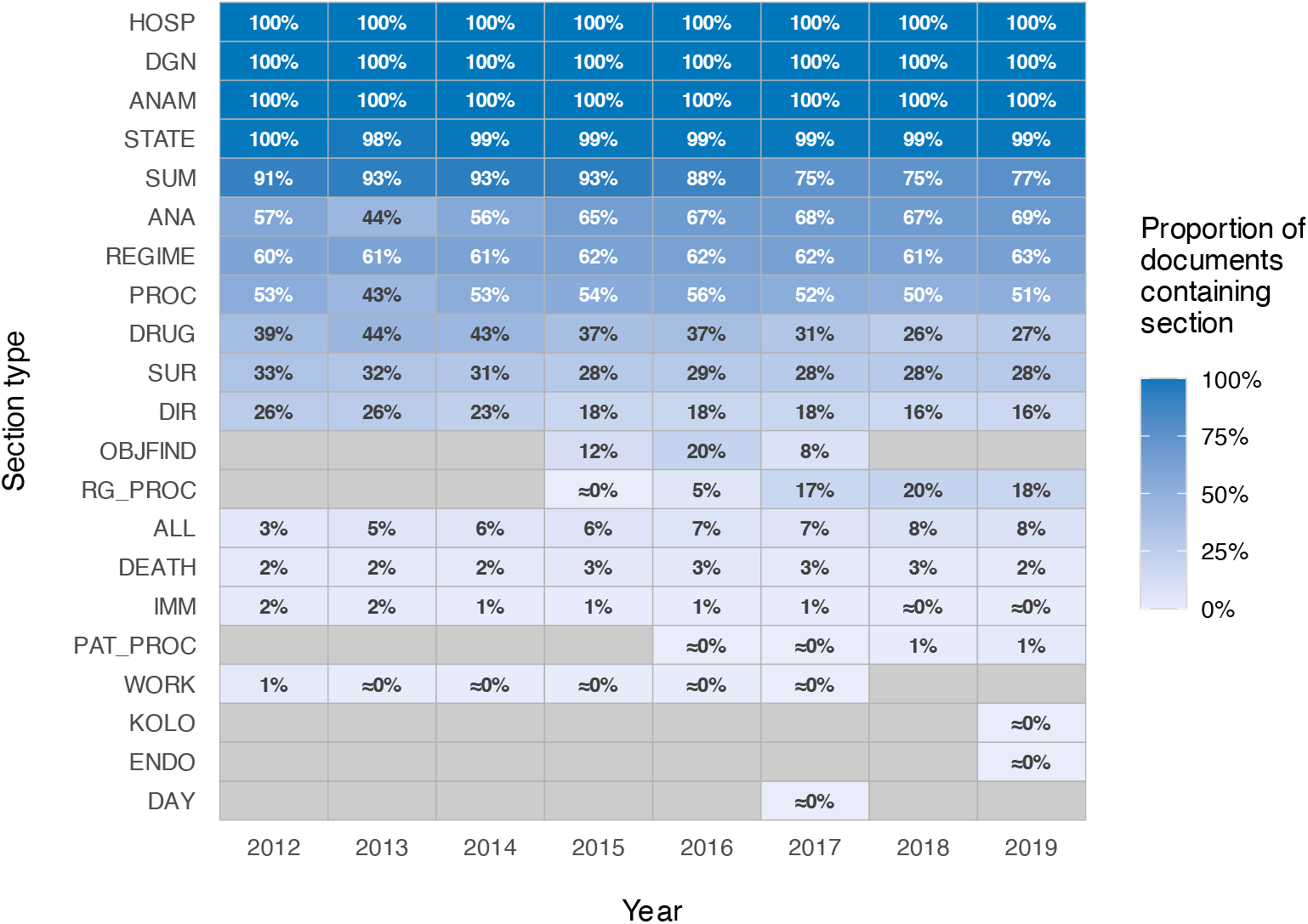
Section presence across years in inpatient discharge summaries.

This composition is not static. `SUM` declined from 91% to 77% between 2012 and 2019, while `ANA` rose from 57% to 69%. New section types entered the landscape: `RG_PROC`, absent before 2015, reached 18% by 2019. Conversely, `DRUG` fell from 39% to 27%, and `WORK` disappeared entirely after 2017. Inpatient summaries thus combine a stable four-section core with a more variable periphery, where both which sections appear and how often they appear changed across 2012–2019.

Analogous heatmaps for the remaining document types (Supplementary Figs. S1–S3) show distinct profiles. Referral response documents underwent the most pronounced shift: `PROC` fell from 100% to 20%, while `ANA` rose from absence to 65% over the same interval. In referrals, `DIR_PURPOSE` and `DIR_SERVICE` appeared from 2017 onward and quickly exceeded 50%. Outpatient discharge summaries had the broadest repertoire (22 types) but with a long tail: only 6 exceeded 30% presence in 2019. Pooled across all four document types and the full 2012–2019 interval, the same skew holds at corpus level: a small subset of section types accounts for the bulk of all section occurrences (Supplementary Fig. S4).

#### Within-section location usage

Structural change also occurs *within* sections. Figure 4 illustrates this using the `ANA` (laboratory investigations) section in inpatient discharge summaries, plotting the mean number of occurrences of each XPath location per section instance.

**Figure 4.**
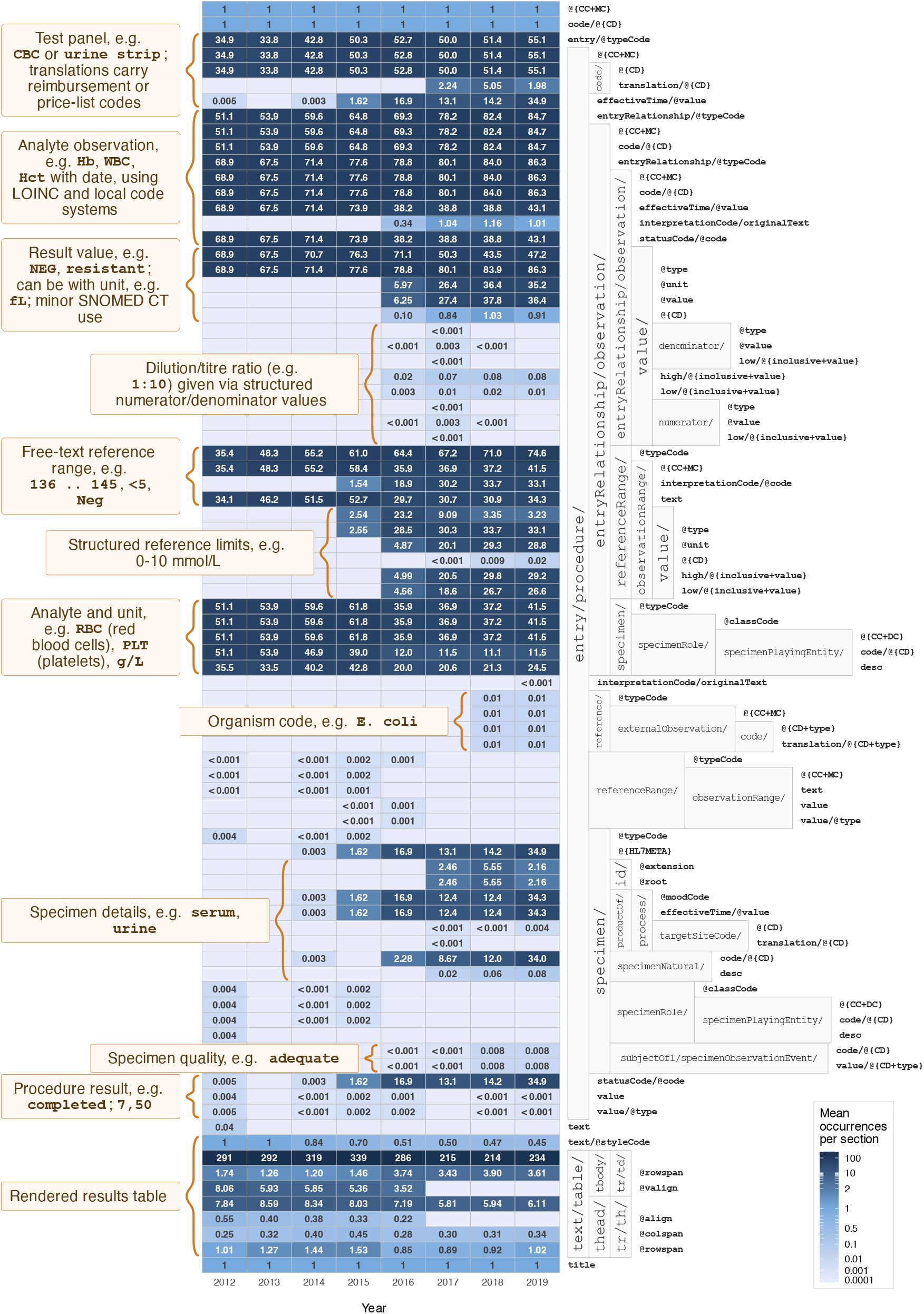
Year-specific mean XPath occurrence within the laboratory investigations (ANA) section of inpatient discharge summaries. Columns denote years; heatmap rows are aligned to XML locations shown in the adjacent partial XPath containment hierarchy. Cell labels give the mean number of occurrences of each location per ANA section, and colour encodes the same quantity via a contrast-enhanced symmetric log-derived transform centred at 1. Rows labelled @{…} are bundles of sibling attributes that co-vary year by year; the bundle legend is given in Supplementary Table S2.

Content density grew: the mean number of entry elements per `ANA` section rose from 35 in 2012 to 55 in 2019 (58% increase), indicating more laboratory test groups per section instance. Alongside this growth, new locations appeared. Procedure-level timestamps (entry/procedure/effectiveTime/@value), nearly absent before 2015 (mean < 0.01), reached a mean of 35 per section by 2019. Structured reference-range boundary paths (e.g. observationRange/value/ high/@{inclusive+value}), absent before 2016, reached means of 27–29 per section by 2019, appearing alongside the pre-existing free-text `observationRange/text` path which remained stable at approximately 34 mean occurrences per section throughout. Similarly, typed observation result paths (observation/value/@value, observation/value/@unit) grew from absence to means of 35–36, while the untyped `observation/value` declined from 69 to 47. Analogous XPath-level heatmaps for the `DGN`, `HOSP`, and `PROC` sections of inpatient discharge summaries are provided in Supplementary Figs. S6–S8. In short, within-section change has two modes: established locations grow denser, and new structured paths emerge—sometimes alongside, sometimes displacing, their untyped predecessors.

Together, these section-level and location-level patterns show that CDA document structure shifted throughout 2012–2019 at every scale examined: which sections appear, how often each appears, and how each is filled.

### Increasing information diversity

Complexity increases over time across several dimensions: value diversity at fixed locations, the number of code systems in use, the number of section types observed, and the cumulative number of unique XPath locations.

#### Value diversity across locations

Across all documents, we identified 892 distinct data locations. Value diversity among these locations is sharply skewed: nearly half (398, 44.6%) always carry a single fixed value, and another 2% are *near-constant* – they collapse to a single value after a light cleaning pass for encoding artefacts, whitespace, and minor label variants. Together, these (near-)constant locations cover 46.6% of locations and 36.2% of all observed field occurrences. Most of the strict-constant share is structural rather than clinical: the HL7 CDA standard mandates fixed values for type, mood, and relationship codes (e.g. @moodCode = EVN, @typeCode = AUT, @classCode = ASSIGNED), so their information lies in the location’s presence, not its value. The near-constant deviations are likewise non-clinical – dominated by encoding errors and minor label drift rather than genuine value diversity. Low value diversity does not, however, imply low clinical relevance: beyond these (near-)constant locations, many fields carry only a handful of values where a small vocabulary is inherent to the clinical domain.

Genuinely high-diversity locations are a small minority, dominated by narrative text fields, patient and document identifiers, and timestamps.

For instance, free-text paragraph content within clinical sections contained over 11.6 million distinct values across roughly 20.8 million occurrences (56% unique), whereas a structural attribute such as the entry type code carried a single fixed value across more than 31.8 million occurrences.

#### Growth in coding systems

The number of unique code systems grew across all document types (Figure 5a). Across all four types combined, the count rose from 45 in 2012 to a peak of 83 in 2018, before declining slightly to 79 in 2019. Growth rates varied substantially by document type: outpatient discharge summaries increased from 40 to 70 code systems (1.8×), inpatient discharge summaries from 33 to 53 (1.6×), referrals from 18 to 40 (2.2×), and referral responses from 13 to 46 (3.5×). The steepest relative growth occurred in referral-related documents, suggesting that structured coding adoption expanded most rapidly in these simpler document types.

**Figure 5.**
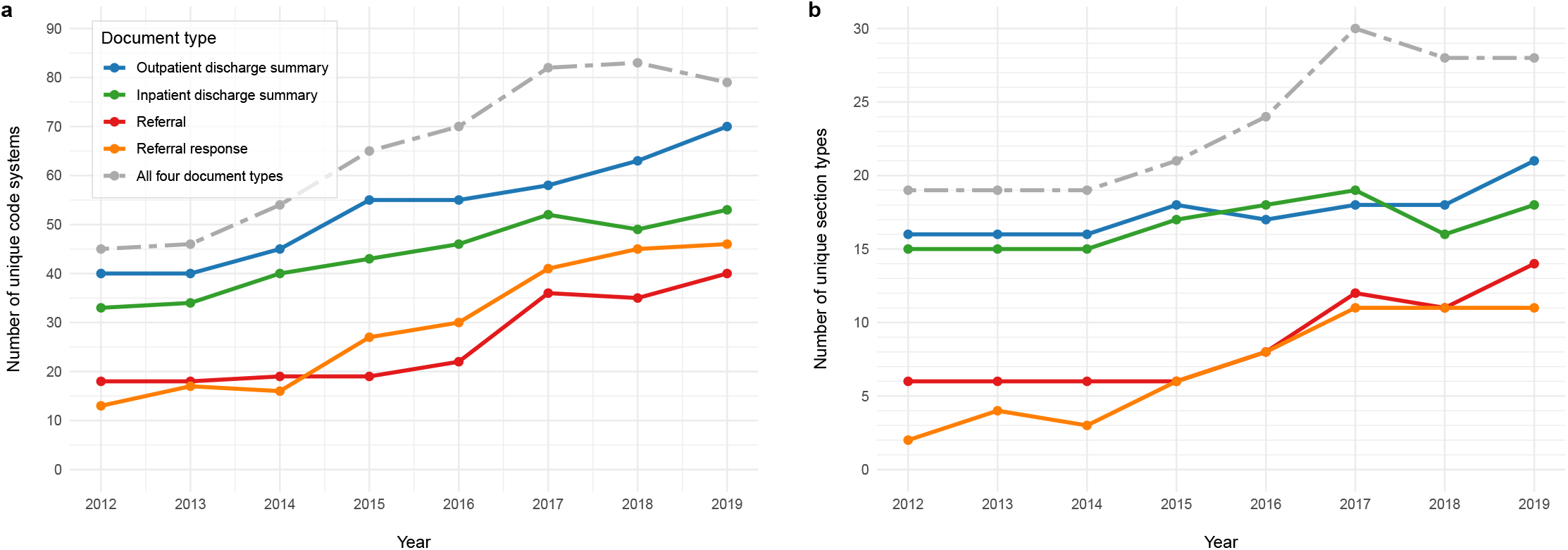
Increasing information diversity over time. **a**, Number of unique code systems by year and document type. **b**, Number of unique section types by year and document type. Colour and line style denote document type; the key is shown in **a**.

#### Growth in section types

Section-type diversity is bounded by the nationally mandated standard but still evolves within the observation window (Figure 5b). Across all four document types, the number of distinct section types rose from 19 in 2012 to 28 in 2019, peaking at 30 in 2017. Growth was unevenly distributed: discharge summaries (both outpatient and inpatient) began with 15–16 section types and grew modestly to 18–21, whereas referral responses expanded from 2 to 11 section types and referrals from 6 to 14, representing roughly five-fold and two-fold increases, respectively. This pattern mirrors the coding-system growth described above, where referral-related documents likewise showed the steepest relative increases, suggesting that simpler, more recently adopted document types absorbed both structural and terminological complexity faster than established ones. The counts include nested sections as well as top-level sections.

#### Growth in unique value locations

Cumulative unique XPath counts within sections grew across all four document types between 2012 and 2019 (Figure 6): outpatient discharge summaries rose from 242 to 522 distinct locations (2.2×), inpatient from 213 to 405 (1.9×), referrals from 114 to 378 (3.3×), and referral responses from 82 to 395 (4.8×), continuing the pattern of steepest relative growth in referral-related documents. Of the 435 new XPaths observed between 2013 and 2019, 44% originated from newly introduced section types and 56% from within pre-existing sections, but this split varied markedly by document type: referral responses derived 74% of their new XPaths from new section types, whereas inpatient discharge summaries drew 76% from pre-existing sections. Thus, the extraction surface expanded through two mechanisms rather than a uniform increase: new section types brought new locations in referral responses, whereas inpatient discharge summaries mainly gained additional locations inside section types already in use.

**Figure 6.**
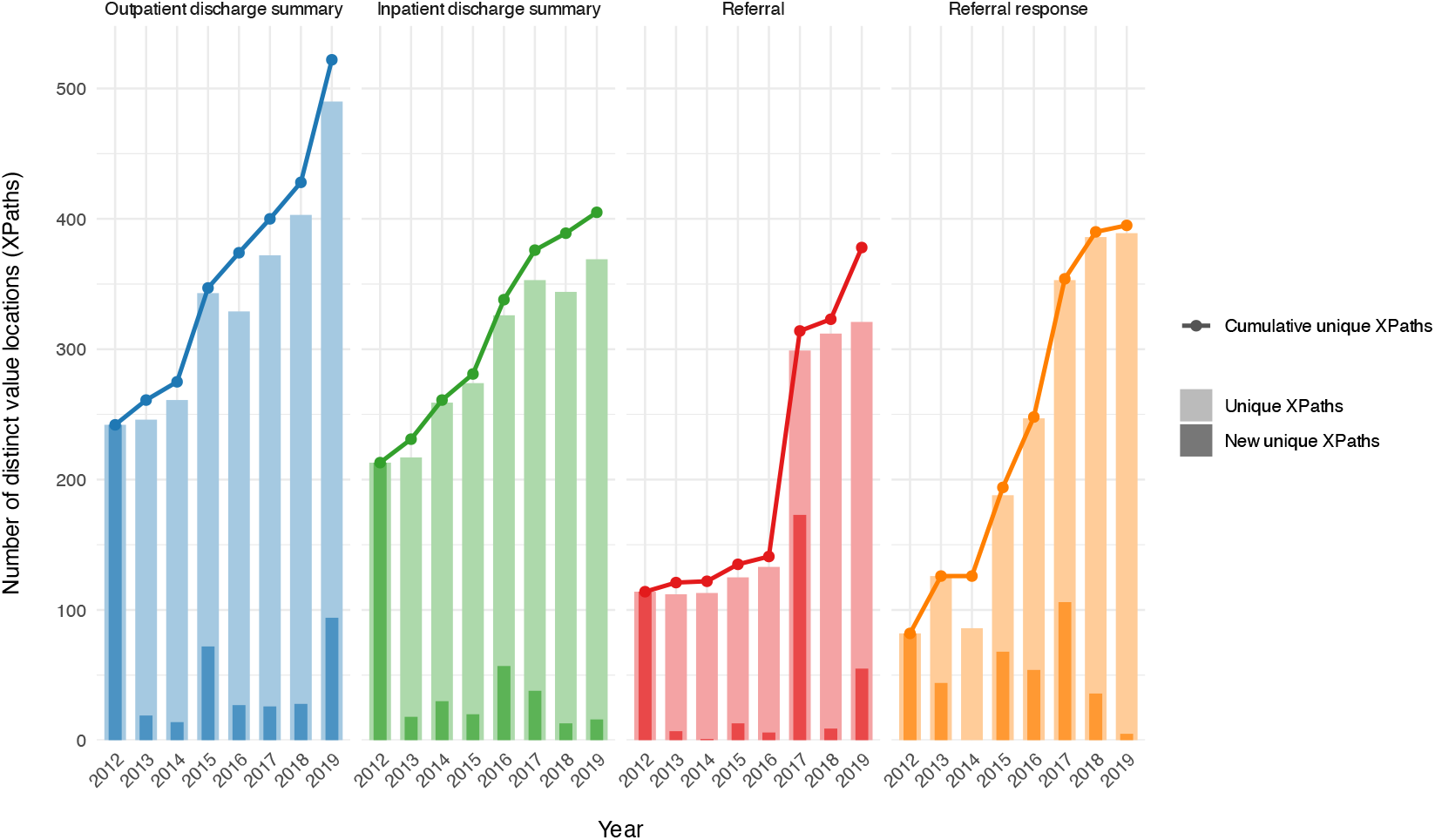
Annual and cumulative counts of distinct value locations (XPaths) within clinical document sections, by document type (2012–2019). Light bars show the number of unique XPaths observed per year; dark overlays indicate XPaths not recorded in any prior year. Lines trace the cumulative total of distinct XPaths over time.

### Standard version uptake

Multiple document versions coexisted in production throughout the observation period: version replacement typically unfolds over years rather than months, with substantial variation across versions and document types. We illustrate this with inpatient discharge summaries and report cross-type patterns for outpatient summaries and referral responses; referrals are omitted because their multiple document subtypes make aggregate version-level analysis impractical.

#### Version distribution

For inpatient discharge summaries, the monthly organisation-level version distribution shows persistent multi-version coexistence rather than sequential displacement (Figure 7). At the start of observation (January 2012), 25 organisations already used two concurrent versions (v2 and v3, both specified before the observation window); by December 2019, six distinct versions were in simultaneous use among 36 organisations, with v7 the most prevalent (16 organisations, 44%), followed by v8 (10, 28%) and v6 (7, 19%). Two specified versions left no trace in our data: v4 (effective July 2014) was never adopted for this document type, and v9 (effective May 2019) reached the observation window too late for any adoption to be assessed.

**Figure 7.**
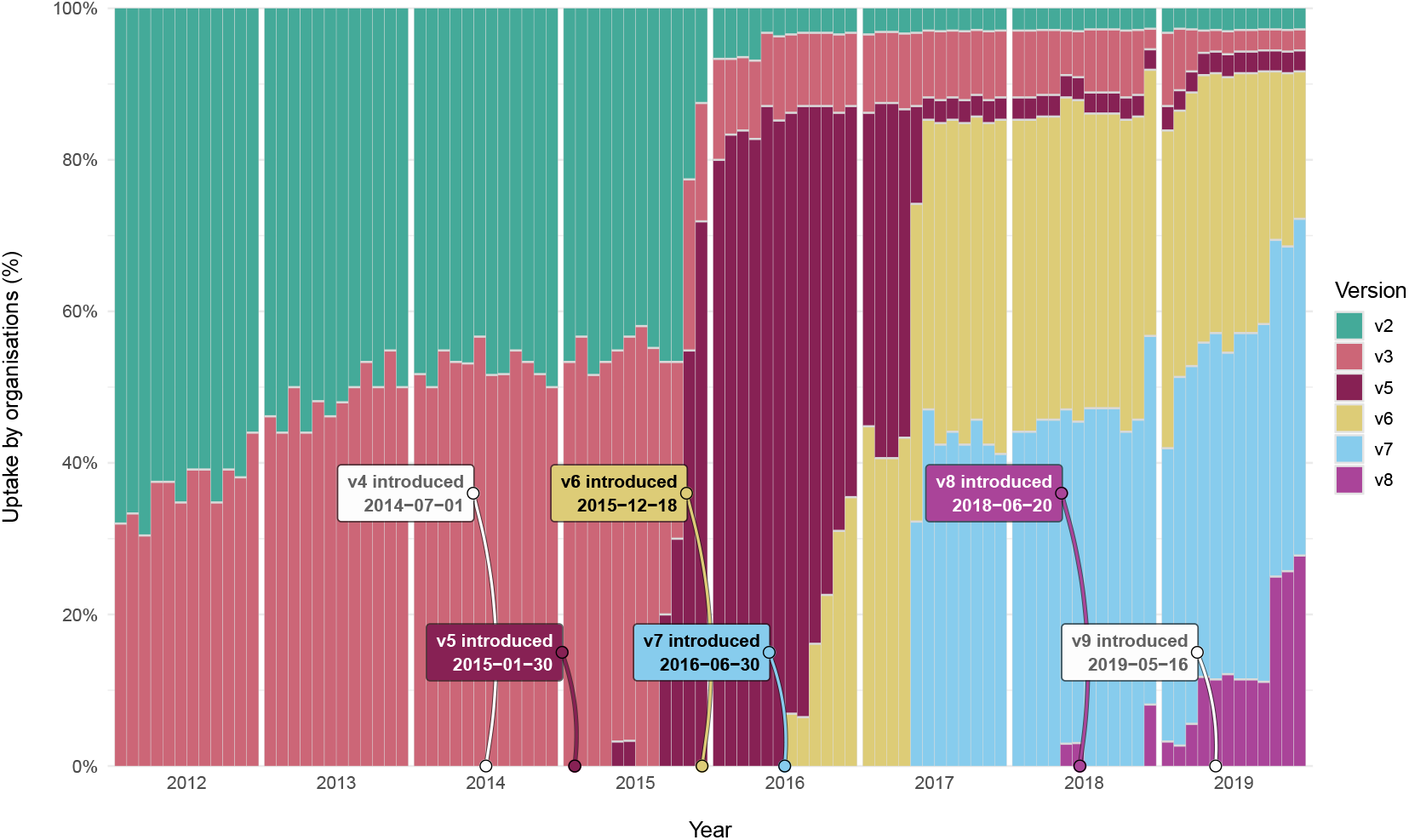
Monthly distribution of organisations across exact document versions for inpatient discharge summaries.

#### Adoption trajectories

Cumulative adoption trajectories, where “≥ v*X*” denotes the proportion of organisations using version *X* or any later version, confirm that version transitions unfold over years (Figure 8). Of the six inpatient versions tracked, three reached 80% organisational adoption during follow-up (at 11, 17, and 78 months after the respective effective date); two later releases (≥ v7, ≥ v8) had not reached it by the end of observation, and one (≥ v2) was already above the threshold when observation began. A Turnbull nonparametric survival estimate [8] over these six versions yields a median time to 80% adoption of 44 months (95% CI: 11–78). The wide confidence interval reflects genuine between-version heterogeneity rather than estimator imprecision: the three observed crossing times span from 11 to 78 months, nearly an order of magnitude. Referral responses showed faster adoption (three exact crossings at 10, 16, and 33 months), whereas most outpatient discharge summary versions introduced during the observation period did not reach the 80% threshold during follow-up. Across document types, version adoption is both heterogeneous within types and type-dependent in pace: multi-year coexistence of versions is the rule rather than the exception within the studied window.

**Figure 8.**
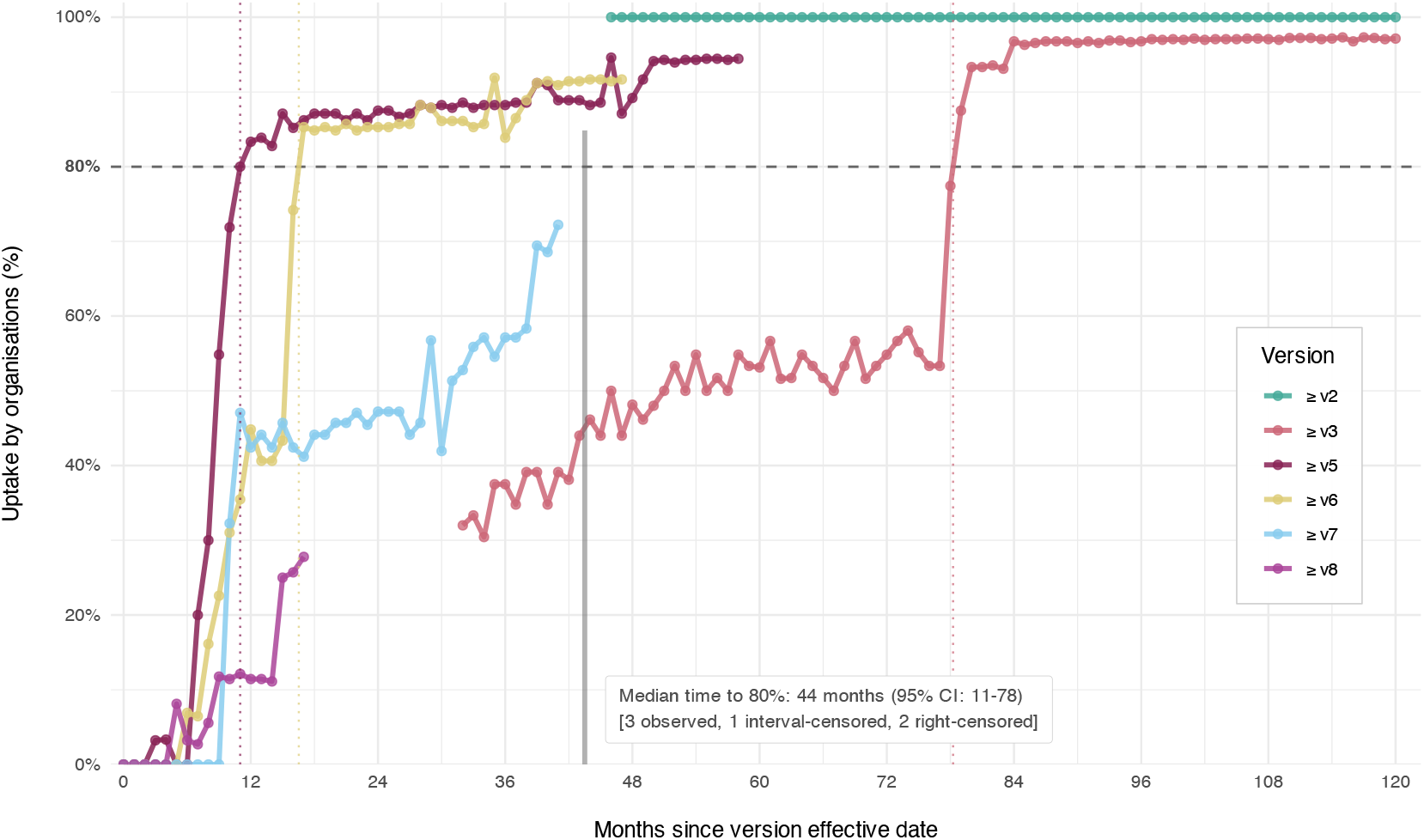
Cumulative organisational adoption trajectories for inpatient discharge summary versions, aligned to version effective dates and showing the 80% uptake threshold. For example, “≥ v6” denotes adoption of version 6 or any later version.

## Discussion

No prior study, to our knowledge, has jointly decomposed clinical document growth in a national CDA-based system across the four concurrent axes used here: volume, per-document size, structural complexity, and standard version uptake. The joint view matters because the dominant growth axis differs by document type, so any single-dimension metric, whether document counts, storage volume, or note length, misrepresents the data-burden trajectory that ETL maintainers, secondary-use researchers, and standards bodies have to plan against. The decomposition methodology transfers directly to other national CDA-based repositories, and, with a corresponding redefinition of the aggregation unit, to FHIR-based or similarly structured repositories. Country-specific benchmarks differ, but the structural phenomena reported here are plausibly general: skewed section distributions, gradual schema evolution, and prolonged version coexistence.

Value variability concentrates in a small minority of data locations. Nearly half of all data locations (44.6%) take only one value, and a further 2% are near-constants whose tail values are entirely encoding errors and inconsistent labelling – the variability at these locations is spurious, not substantive. Together these account for 36.2% of all field occurrences when weighted by frequency. Most strict-constant locations are CDA-mandated structural attributes whose information lies in their presence rather than their value; the genuinely variable content, principally narrative text, identifiers, and timestamps, resides in a small minority of locations that nevertheless generate a disproportionate share of occurrences. Separating CDA-mandated structural attributes from clinically meaningful low-cardinality fields would require an explicit location classification this study does not provide, but the skewed distribution itself is directly informative for ETL prioritisation.

The dominant growth driver differed by type: outpatient summaries grew through patient reach and document frequency, inpatient summaries through document size alone, and referral-related documents through every axis simultaneously. The same heterogeneity reappeared at finer scales. New structural locations entered predominantly through pre-existing sections in inpatient summaries, but through entirely new sections in referral responses. Version-uptake pace was equally type-dependent, ranging from under a year for the fastest types to incomplete adoption by the end of the observation window. Within the laboratory section that drove most of the inpatient size growth, structured reference-range boundaries and typed observation results emerged from 2015–2016 alongside, not in place of, the pre-existing free-text paths. ETL pipelines therefore cannot assume a clean migration from one encoding paradigm to another, even within a single section over a few years. Backward-compatibility windows, monitoring cadences, and storage assumptions calibrated for one document type may not hold for another.

Three practical consequences follow for ETL pipeline design and standards governance. First, the skewed section-presence distribution means that a small subset of sections covers most observed content, making selective extraction feasible for narrowly scoped tasks; however, membership of that high-coverage set shifts over time and must be monitored rather than assumed fixed. Sections such as `ANA` and `RG_PROC` rose from low or zero prevalence to substantial presence within the observation window. When future analytic questions are uncertain, a full-document structural inventory is useful as a discovery layer: low-prevalence, nested, and newly emerging data locations remain queryable even before their clinical or analytic relevance has been decided, which reduces dependence on sporadic XML-level inspection when the document structure changes. Second, new section types, new locations within existing sections, and new code systems appear progressively rather than at version boundaries. This gradual-evolution pattern means that monitoring has to cover both section-level and within-section change; selective extraction remains feasible provided it is paired with discovery of unrecognised section types and locations and with graceful degradation when document structure differs from expectations. Third, the long multi-version coexistence periods mean that standards bodies, repository operators, and ETL maintainers need explicit backward-compatibility windows rather than assuming that a new version rapidly displaces its predecessors in routine use. For secondary-use researchers constructing observational databases in standard models such as Observational Medical Outcome Partnership (OMOP) Common Data Model (CDM) [9, 10], applying natural language processing to clinical narratives [11, 12, 13], or training large language models on clinical text [14], the combined growth in document size, section types, value locations, code systems, and coexisting document versions means that the input data surface is both larger and more heterogeneous than document counts alone imply; schema drift over time should therefore be factored into study-period selection and data-quality assessment.

Existing benchmarks each measure one dimension of the joint picture we decompose; cross-checking our per-axis numbers against them validates each axis individually while underlining what single-dimension studies miss. Our per-document size growth (AAPC +1.3% to +5.5%) aligns with the 4.8% CAGR in outpatient note length reported by Rule et al. [1] over 2009–2018, even though that study measured narrative word counts at a single US institution whereas we measure total field-value characters including structured content in a national system. The higher rates we observe for inpatient (+5.5%) and referral-response (+5.0%) types likely reflect structured field expansion, such as new coded entries and reference ranges, rather than narrative length growth alone. Rao et al. [15] documented a complementary pattern in US primary care: visit frequency per capita declined by 20% over 2008–2015 while diagnoses addressed per visit rose by 15%, consistent with our finding that document frequency alone understates the true growth in data volume. Patterson et al. [2] reported approximately 30% CAGR in notes per emergency department encounter over 17 years, but their metric captures cumulative chart burden rather than annual new production. This distinction underscores why per-year decomposition is needed alongside cumulative views. Narrower hospital-only datasets already show growth in administrative coding intensity. In US hospital discharge records from five states, discharges assigned to the highest coded severity tier increased by about 4% per year from 2011 to 2019 [16]; in German hospital records, the mean number of coded comorbidity groups per record rose by about 1%–3% per year from 2005 to 2012 [17]. Our national CDA repository shows a parallel increase in breadth: distinct code systems used across four document types rose from 45 in 2012 to 79 in 2019. Clinical coding is expanding both in coding density within records and in the breadth of vocabularies used. The Finnish Kanta Patient Data Repository [18], the most structurally comparable national system, underwent rapid growth during provider onboarding and exhibited category-specific growth patterns (laboratory data growing tenfold after 2019, diagnoses stabilising after 2017), consistent with the type-dependent heterogeneity we observe.

At the sub-document structural level, comparable evidence is cross-sectional and small in scale. The SMART C-CDA Collaborative analysed 91 documents from 21 EHR technologies and recorded 615 observations of error and encoding variation across vendors [19], and a follow-up of 401 C-CDA 2.1 documents reported the same data element (encounter time) carried in four distinct XPath patterns across vendors [20]. In real exchange traffic, laboratory sections were populated in 9%–81% of documents across nine US eHealth Exchange partners [21]. A recent template-driven CDA-to-OMOP study has called explicitly for future work to investigate the heterogeneity of CDA documents constrained by the same template [22]; this question requires production-scale analysis below the section level.

For version uptake, no direct CDA benchmark exists; proxy comparators converge on similar timescales. The closest analogue is the 2015 update to United States hospital EHR certification criteria, which bundled an upgrade of the Consolidated CDA (C-CDA) from R1.1 to R2.1 onto a near-saturated base (only 3% of hospitals lacked certified technology) [23]. Hospitals took roughly 38 months to cross 80% adoption [24, 25], broadly in line with our 44-month median for inpatient discharge summaries, though that figure measures certification attestation rather than document-level conformance and is backed by federal incentives absent in our setting. The same multi-year pattern appears in other standards, where HL7 v2.5.1 capability took five to seven years to reach most jurisdictions in US childhood immunisation registries [26]. Transitions are rarely clean: within C-CDA itself, prior-template documents still made up about one-fifth of a 2018 certification testing pool [20].

Across the comparison, each axis individually has a published benchmark, but the joint pattern (the same document types ranking differently on different axes) has no published analogue.

Several limitations remain, all tied to scope and measurement choices. The observation window covers 2012–2019, a post-implementation maturation period rather than contemporary practice; the methodology, however, transfers directly to more recent data. The analysis draws on a 10% random sample of the Estonian population (147,819 patients, approximately 5 million documents), adequate for section-level prevalence but liable to underrepresent rare structural elements. Uptake is measured at the document-version level rather than the section level, because section presence is unreliable as an adoption proxy when sections are clinically optional; the version-level proxy in turn conflates targeted adoption with version leapfrogging.

Most longitudinal accounts of EHR document growth and standards uptake track a single axis at a time. The decomposition presented here treats those axes jointly: which are growing, in which document types, on which timescales, and with what version-coexistence overhead. For federated research networks and the secondary-use infrastructure built on standards-based clinical-exchange repositories, what a system will support in three years depends as much on how growth has been distributed across these axes as on its current conformance. That trajectory is only visible through joint multi-axis decomposition of the kind demonstrated here.

## Methods

### Data extraction and storage

The analytical corpus comprises CDA R2 documents generated between 1 January 2012 and 31 December 2019, representing a 10% random sample of the Estonian population. Document-type composition, patient counts, exclusions of malformed or empty documents, and the set of section types are reported with the empirical findings (Tables 1–3).

The analytical workflow follows a full-extraction strategy in which CDA R2 XML documents are transformed by Extensible Stylesheet Language Transformations (XSLT) into per-document comma-separated values (CSV) files and then loaded into the DuckDB analytical database [27]. Each CSV row represents a single extracted value together with its source file path, its sequence number within the file, and an XPath expression, yielding records with fields `file_path`, `seq_no_in_file`, `xpath`, and `value`. The XPath preserves structural context, including namespace-qualified structure and selected semantic qualifiers such as section codes, while the sequence number preserves document order. These rows populate a generic value-level table (`kv_store`). A companion document-level table (`file_info`) is derived from the extracted records by identifying unique document and patient identifiers from file paths and by attaching document-level attributes such as type and document date. The document date is derived from a predefined set of document-level CDA time fields stored in `kv_store`, using a prioritised fallback, and is subsequently reduced to year or year-month as required for temporal analyses. Additional post-processing produces three derived structures used downstream. A normalised XPath form removes namespace prefixes to yield a canonical structural representation, supporting unique location counts that are not confounded by alias variation. An organisation table groups documents by submitting organisation, identified from document headers, and supports the version uptake analysis. A code system table maps observed terminology identifiers to canonical OIDs – consolidating equivalent variants, such as those differing only in version su?x or in provider-specific intermediate segments – and supports the code system diversity analysis.

Full extraction is necessary for the analyses reported in this study: computing document and section size requires summing all extracted values, while unique location counts and code system diversity depend on discovering – rather than pre-specifying – the structural locations and code system identifiers present in the corpus. Selective parsing, which commits to a target schema before the data are fully understood, supports neither.

### Analytical measures and statistical methods

The analysis is organised as a decomposition of longitudinal change rather than as a single aggregate growth estimate. Measures are derived at multiple levels, including document counts, section occurrence, section-level content volume, distinct structural locations (XPaths), code-system diversity, and organisation-level document-version uptake.

Temporal trends in annual counts and sizes are summarised using average annual percent change (AAPC) with 95% Wald confidence intervals. AAPC is used for the main trend summaries because it provides an interpretable annualised measure together with uncertainty quantification over the full observation period.

Section occurrence is quantified as the proportion of documents containing a given section type within a document type and year. Section-type diversity is measured as the number of distinct section codes observed per year. Code-system diversity is measured as the number of unique canonical code systems observed per year, combining header-level and section-level coding locations.

Structural diversification within sections is assessed from distinct XPath locations. For each document type and year, we count the number of unique XPaths observed and track their cumulative growth over time. To distinguish expansion through new section types from expansion within existing section types, we classify each newly observed XPath by the section types in which it first appears. An XPath is considered to arise within a pre-existing section if, in its first observed year, it appears in any section type already observed in any earlier year; otherwise it is classified as originating from a newly introduced section type.

Growth in document size is further decomposed by section, separately within each document type. For each section *s* in year *t*, we sum the character lengths of all values extracted from the section, per document, and take the median across documents to obtain the annual section size *M*_*s,t*_. A section’s absolute contribution to growth is the first-to-last change in this measure, 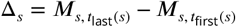, where *t*_first_(*s*) and *t*_last_(*s*) are the first and last years in which the section is observed (a per-section window, not necessarily 2012–2019). The net document-size change for that document type is the sum of all sections’ contributions, Δ_total_ = ∑_*s*_ Δ_*s*_, and the percentage share attributable to section *s* is 100 ⋅ Δ_*s*_/Δ_total_.

Document-version uptake is analysed at the organisation level using the document standard version as the uptake proxy. This proxy is preferred to section-level presence because many sections are clinically optional, so their absence does not necessarily indicate non-adoption. For each version, time is aligned to the version’s effective date, and cumulative organisational adoption is defined as the proportion of organisations using “≥ v*X*”, that is, version *X* or any later version. Because observed monthly percentages can fluctuate owing to changes in the number of contributing organisations, per-version adoption trajectories are smoothed with isotonic regression [28, 29] to recover the underlying monotone adoption signal. Time to 80% adoption is then classified as exact, interval-censored, or right-censored according to whether the threshold crossing is directly observed, known only to have occurred before first observation or between observations, or not reached during follow-up [30]. Median time to 80% adoption and 95% confidence intervals are estimated with the Turnbull nonparametric maximum likelihood estimator for interval-censored data [8], using the `survival` package in R [31, 32].

### Ethics approval and consent to participate

The study was approved by the Ethics Committee of the University of Tartu (No. 300/T-23) on 20 January 2020 and by the Estonian Bioethics and Human Research Council (No. 1.1-12/653) on 14 April 2020; extensions were subsequently granted by the Estonian Bioethics and Human Research Council (No. 1.1-12/613) on 4 March 2025 and by the Ethics Committee of the University of Tartu (No. 401/T-34) on 19 May 2025. The requirement for individual informed consent was waived by the approving committees, as the study used pseudonymised retrospective data from existing national health databases.

## Acknowledgements

This work was supported by the Estonian Research Council (PRG1844). The study was funded by the European Union and co-funded by the Ministry of Education and Research (TEM-TA72). The European Union funded the project under its Horizon Europe research and innovation programme (grant agreement No 101060011, TeamPerMed) and co-funded the research through the European Regional Development Fund (Project No 2021-2027.1.01.24-0444). Views and opinions expressed are, however, those of the authors only and do not necessarily reflect those of the European Union or the European Research Executive Agency. Neither the European Union nor the granting authority can be held responsible for them. This work was also supported by the Estonian Centre of Excellence in Artificial Intelligence (EXAI), funded by the Estonian Ministry of Education and Research grant TK213, as well as by STACC’s regular base funding for R&D from the Ministry of Education and Research.

We thank Sirli Tamm, Marek Oja, and Raivo Kolde for constructive feedback on earlier versions of this work.

Frontier large language models were used to assist with code debugging and to improve the grammar and fluency of the manuscript. All AI-generated suggestions were reviewed and verified by the authors, who bear full responsibility for the content.

The data processing described in this work was carried out at the High Performance Computing Center of the University of Tartu.

## Declarations

### Ethics approval

The study was approved by the Ethics Committee of the University of Tartu (No. 300/T-23) on 20 January 2020 and by the Estonian Bioethics and Human Research Council (No. 1.1-12/653) on 14 April 2020; extensions were subsequently granted by the Estonian Bioethics and Human Research Council (No. 1.1-12/613) on 4 March 2025 and by the Ethics Committee of the University of Tartu (No. 401/T-34) on 19 May 2025.

### Data availability

The data underlying this work cannot be shared publicly for the privacy of individuals who participated in the study. The data were obtained from national health databases and can be requested via the Estonian Research Ethics Committee. Auxiliary metadata used during the study came from TEHIK’s Information Centre, the MEDRE registry of the Estonian Health Board, and the e-Business Register.

### Code availability

The code underlying this study is not publicly available at the time of preprint posting. A reviewer-only archive containing the principal source-code components and accompanying documentation for data processing, transformation, and analysis has been provided to the journal for editorial and peer-review assessment. The archive contains no patient-level data. Public release is deferred because the code was developed for a restricted analytical environment and requires institutional disclosure review before public distribution. Upon acceptance and before publication, the authors will make a disclosure-reviewed version of the code publicly available in a persistent repository and will update this statement with the repository link and/or DOI.

### Author contributions

H.-A.T.: Conceptualisation, Methodology, Software, Formal Analysis, Data Curation, Validation, Visualisation, Writing - Original Draft. S.R.: Conceptualisation, Methodology, Supervision, Writing - Review and Editing. S.L.: Conceptualisation, Methodology, Formal Analysis, Writing - Review and Editing. J.V.: Writing - Review and Editing, Funding Acquisition.

