## Supplementary information for "Decomposing growth in a national HL7 CDA clinical document repository"

*medRχiv preprint supplement — see published version for the version of record once available*

This supplement provides extended visualisations and a complete section-type catalogue beyond what is presented in the main article. Figures S1–S4 extend the section-presence and section-skew analyses to the remaining three document types; Figures S6–S8 extend the within-section XPath analysis to further inpatient discharge summary sections. Supplementary Table S1 and Figure S5 catalogue the 125 section types defined in the Estonian CDA R2 specification and place them on an introduction timeline, situating the four studied document types within the wider Estonian section-type landscape and identifying additional material to which the same decomposition approach could be extended. Supplementary Table S2 documents the XPath compaction notation used in the within-section heatmaps.

#### Contents

**1 Section presence across years for the remaining document types (Figures S1–S3)** 2

**2 Cumulative distribution of sections (Figure S4)** 4

**3 Complete section type catalogue (Table S1)** 4

**4 Section type introduction timeline (Figure S5)** 11

**5 XPath compaction notation for within-section heatmaps (Table S2)** 12

**6 Additional XPath-level heatmaps for inpatient discharge summaries (Figures S6–S8)** 13

### 1 Section presence across years for the remaining document types (Figures S1–S3)

Figures S1–S3 complement the main-text inpatient section-presence heatmap by showing the same analysis for the three remaining document types: outpatient discharge summaries (Figure S1), referrals (Figure S2), and referral responses (Figure S3). The same colour scale, axis structure, and annotation style are used to support direct visual comparison.

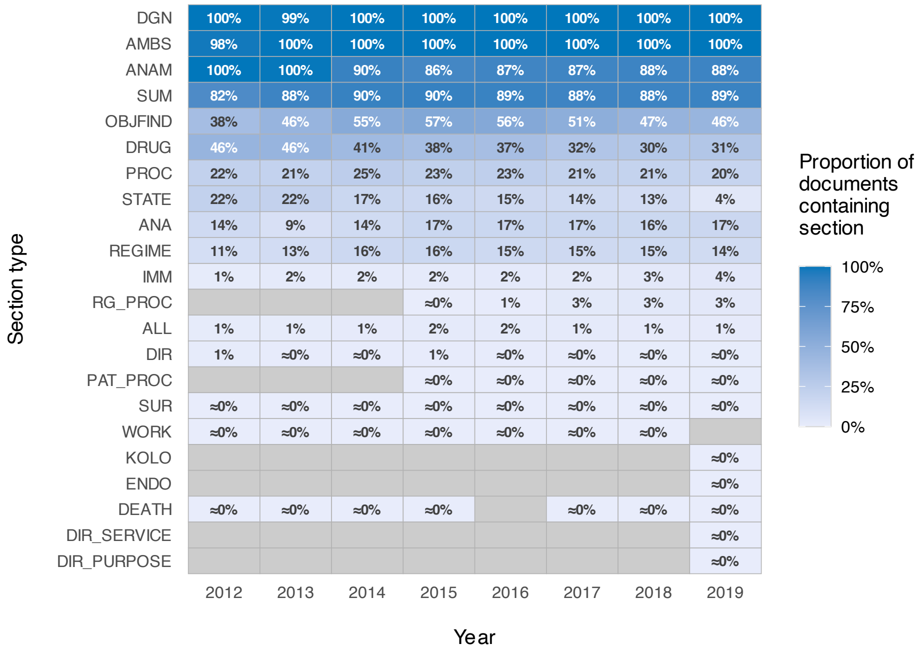


**Figure S1.** Section presence across years in outpatient discharge summaries.

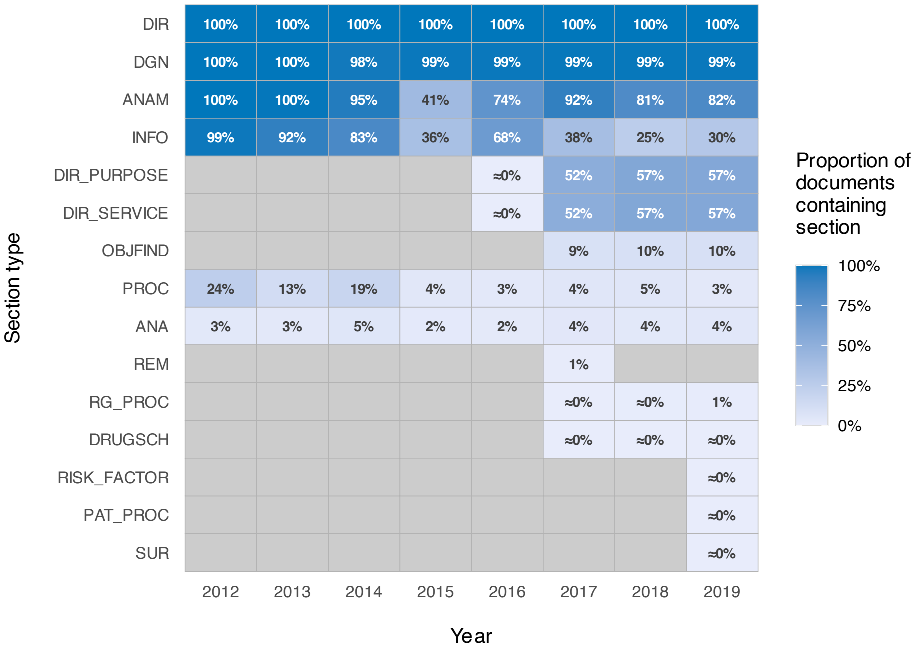


**Figure S2.** Section presence across years in referrals.

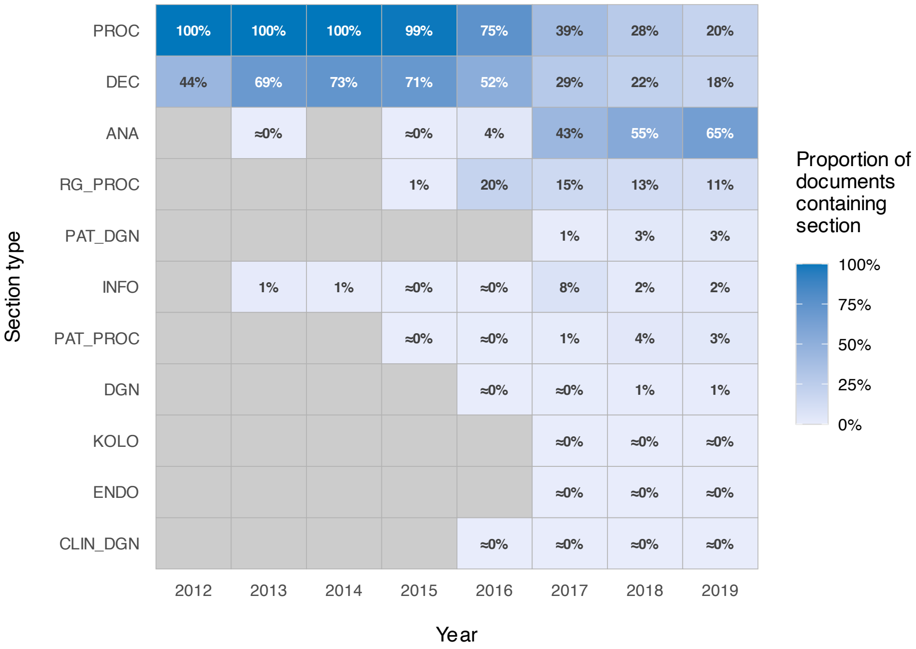


**Figure S3.** Section presence across years in referral responses.

#### 2 Cumulative distribution of sections (Figure S4)

Figure S4 provides a single aggregate view of section skew across all years and document types. It is intended to complement the main-text inpatient section-presence heatmap and the analogous heatmaps for the remaining three document types (Figs. S1–S3) by showing directly that a small number of section types account for the majority of section occurrences. Of the 31 section types observed across the four document types, the 13 most prevalent together account for approximately 95% of all section occurrences in the pooled corpus, with the remaining 18 types contributing the residual 5% long tail.

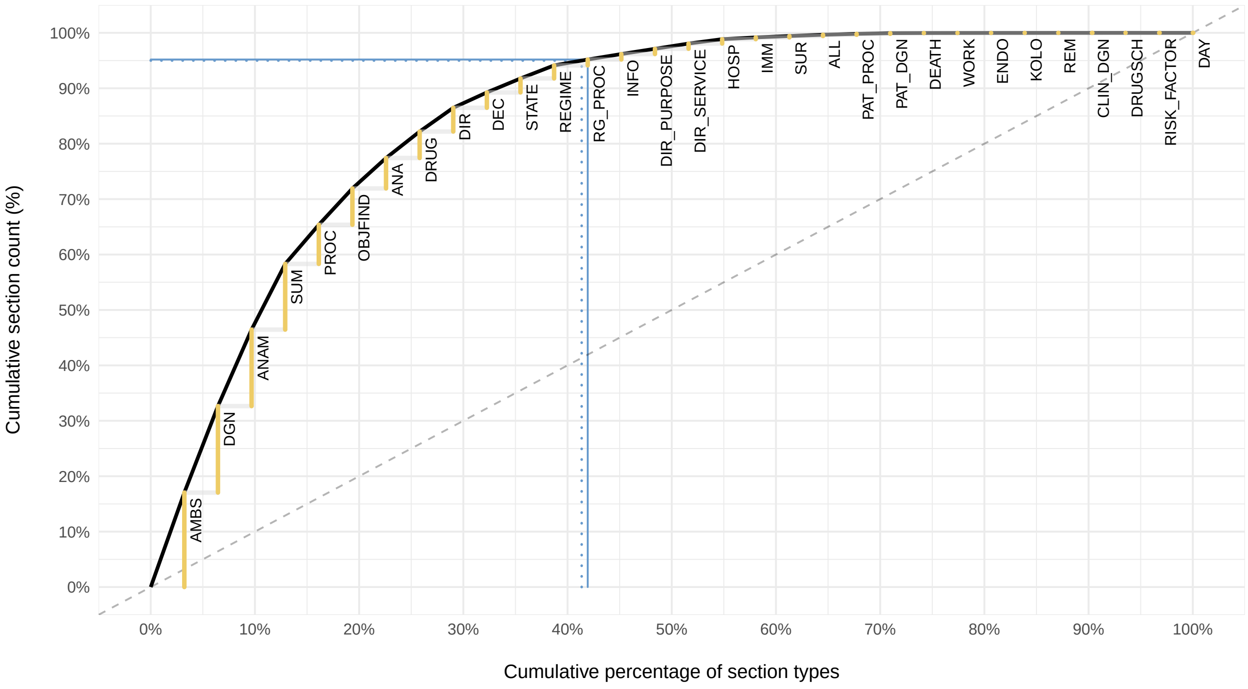


**Figure S4.** Cumulative distribution of section counts across the analysed corpus. Section types ordered by descending contribution. Dotted blue reference lines mark the 95% cumulative-coverage threshold; solid blue reference lines mark the first cumulative point at or above this threshold, reached after the 13th most-prevalent section type.

#### 3 Complete section type catalogue (Table S1)

Table S1 lists the full set of 125 section types tracked from the Estonian CDA R2 specification source used during article preparation. The "Observed in dataset" column distinguishes section types present in the four analysed document types from section types that are part of the broader specification landscape but absent from the 2012–2019 study corpus.

**Table S1.** Complete section type catalogue. Section names are given in English, followed by the Estonian term in italics. In the "Observed in dataset" column, ✓ denotes section types observed in the study dataset and – denotes section types not observed.

| Section code | Section name | Valid since | Observed in dataset |
|---|---|---|---|
| AGE | Age<br>*vanus* | 2007-09-07 | – |
| ALL | Allergy<br>*allergia* | 2007-09-07 | ✓ |
| AMBS | Outpatient case<br>*ambulatoorne haigusjuhtum* | 2007-09-07 | ✓ |
| ANA | Laboratory investigations<br>*laboratoorsed uuringud* | 2007-09-07 | ✓ |
| ANAM | Anamnesis, diagnosis justification and disease course<br>*anamnees, diagnoosi põhjendus ja haiguse kulg* | 2007-09-07 | ✓ |
| BANA | Newborn analyses<br>*vastsündinu analüüsid* | 2007-09-07 | – |
| BHOSP | Hospital stay<br>*haiglas viibimine* | 2007-09-07 | – |
| BIRTH | Newborn birth data<br>*vastsündinu sünniandmed* | 2007-09-07 | – |
| BLEAVESTATE | Newborn condition at discharge<br>*vastsündinu seisund haiglast väljakirjutamisel* | 2007-09-07 | – |
| BLRH | Blood group / Rf<br>*veregrupp / Rf* | 2007-09-07 | – |
| BSTATE | Newborn health status assessment<br>*vastsündinu tervisliku seisundi hinnang* | 2007-09-07 | – |
| CASELST | Last visit or hospitalization<br>*viimane visiit või hospitaliseerimine* | 2007-09-07 | – |
| CIND | Immunization contraindication<br>*immuniseerimise vastunäidustus* | 2007-09-07 | – |
| CUSTOM | Health habits<br>*terviseharjumused* | 2007-09-07 | – |
| DAY | Day care case<br>*päevaravi haigusjuhtum* | 2007-09-07 | ✓ |
| DEATH | Death<br>*surm* | 2007-09-07 | ✓ |
| DGN | Final clinical diagnosis<br>*lõplik kliiniline diagnoos* | 2007-09-07 | ✓ |

| Section code | Section name | Valid since | Observed in dataset |
|---|---|---|---|
| DGNCHR | Chronic disease<br>*krooniline haigus* | 2007-09-07 | – |
| DGNVIR | Viral and infectious diseases<br>*viirus- ja nakkushaigused* | 2007-09-07 | – |
| DIR | Referral<br>*vastuvõtule pöördumine* | 2007-09-07 | ✓ |
| DISABL | Disability<br>*puue* | 2007-09-07 | – |
| DOC | Issued documents<br>*väljastatud dokumendid* | 2007-09-07 | – |
| DRUG | Prescribed and dispensed medications<br>*väljakirjutatud ja väljastatud ravimid* | 2007-09-07 | ✓ |
| DRUGLST | Medications purchased in the last 3 months<br>*ravimid välja ostetud viimase 3 kuu jooksul* | 2007-09-07 | – |
| FAMILY | Family situation<br>*pere olukord* | 2007-09-07 | – |
| GROWTH | Growth data<br>*kasvu andmed* | 2007-09-07 | – |
| GUID | Counseling<br>*nõustamine* | 2007-09-07 | – |
| HHANAM | Nursing anamnesis<br>*õendusanamnees* | 2007-09-07 | – |
| HHPROC | Performed procedures<br>*teostatud toimingud* | 2007-09-07 | – |
| HOSP | Hospital stay<br>*haiglas viibimine* | 2007-09-07 | ✓ |
| IMM | Immunization<br>*immuniseerimine* | 2007-09-07 | ✓ |
| INFO | Additional information about the patient<br>*lisateave patsiendi kohta* | 2007-09-07 | ✓ |
| MANA | Mother's analyses<br>*ema analüüsid* | 2007-09-07 | – |
| MANTOUX | Tuberculin/Mantoux test<br>*Tuberkuliin/Mantoux test* | 2007-09-07 | – |
| MENTAL | Mental background and development<br>*vaimne taust ja areng* | 2007-09-07 | – |
| OBJFIND | Objective finding<br>*objektiivne leid* | 2007-09-07 | ✓ |
| PHYS | Examination<br>*läbivaatus* | 2007-09-07 | – |

| Section code | Section name | Valid since | Observed in dataset |
|---|---|---|---|
| PRGN | Pregnancy<br>*rasedus* | 2007-09-07 | – |
| PROC | Examinations and procedures<br>*uuringud ja protseduurid* | 2007-09-07 | ✓ |
| PROCLST | Surgical procedures in the last month<br>*kirurgilised protseduurid viimase kuu jooksul* | 2007-09-07 | – |
| PROG | Development assessment<br>*arengu hindamine* | 2007-09-07 | – |
| PSYHSOC | Psychosocial background and development<br>*psühhosotsiaalne taust ja areng* | 2007-09-07 | – |
| REGIME | Regimen and treatment recommendations<br>*režiimi ja ravialased soovitused* | 2007-09-07 | ✓ |
| REM | Remarks<br>*märkused* | 2007-09-07 | ✓ |
| SPEC | Special arrangements<br>*erikorraldused* | 2007-09-07 | – |
| STATE | Condition at discharge<br>*seisund väljakirjutamisel* | 2007-09-07 | ✓ |
| STATPROC | Performed procedures<br>*teostatud toimingud* | 2007-09-07 | – |
| STPROC | Significant surgical procedures<br>*olulisemad kirurgilised protseduurid* | 2007-09-07 | – |
| SUM | Summary of patient treatment<br>*kokkuvõte patsiendi ravist* | 2007-09-07 | ✓ |
| SUR | Operations<br>*operatsioonid* | 2007-09-07 | ✓ |
| TELCONSULT | Participation in telemedicine consultation<br>*telemeditsiinilisel konsultatsioonil osalemine* | 2007-09-07 | – |
| TRUST | Extent of authorization<br>*volituse ulatus* | 2007-09-07 | – |
| WORK | Work environment and arrangement changes<br>*töökeskkonna ja -korralduse muutmine* | 2007-09-07 | ✓ |
| PVIIT | Image reference<br>*pildiviit* | 2008-02-19 | – |
| WILL | Declaration of will for healthcare provision<br>*tahteavaldus tervishoiuteenuse osutamiseks* | 2008-06-05 | – |
| CONSULT | Specialist consultations<br>*Eriarstide konsultatsioonid* | 2009-03-25 | – |

| Section code | Section name | Valid since | Observed in dataset |
| --- | --- | --- | --- |
| PASTPRGN | Previous pregnancies/births data (based on statements)<br>*Eelmiste raseduste / sünnituste andmed (ütluspõhine)* | 2009-03-25 | – |
| PELVIS_PROC | Vaginal examination<br>*Vaginaalne vaatlus* | 2009-03-25 | – |
| PRE | Prescription<br>*retsept* | 2009-01-15 | – |
| PRGN_DL | Delivery summary and/or maternal inpatient summary<br>*Sünniepikriis ja/või ema statsionaarne epikriis (DL)* | 2009-03-25 | – |
| RAV_PROC | Gravidogram<br>*Gravidogramm* | 2009-03-25 | – |
| ALLPROC | Examinations, analyses, operations<br>*Uuringud, analüüsid, operatsioonid* | 2012-07-31 | – |
| AMBNEED | Crew assessment of ambulance necessity<br>*Brigaadi hinnang juhtumile kiirabi vajalikkusele* | 2012-07-31 | – |
| AMBUBU | Assisted another ambulance crew<br>*Oldi abis teisel kiirabibrigaadil* | 2012-07-31 | – |
| AMBULANCEDRUG | Ambulance-administered medications<br>*Kiirabi manustatud ravimid* | 2012-07-31 | – |
| AMBULANCEPROC | Ambulance procedures<br>*Kiirabi teostatud protseduurid* | 2012-07-31 | – |
| AMBVISITSUMMARY | Ambulance visit outcome<br>*Kiirabi visiidi tulemus* | 2012-07-31 | – |
| ANAMEC | EMS card anamnesis<br>*Kiirabikaardi anamnees* | 2012-07-31 | – |
| CPR | Cardiopulmonary resuscitation<br>*Elustamine* | 2012-07-31 | – |
| DRUGLST6 | Prescriptions or administered medications in the last 9 months<br>*Viimase 9 kuu jooksul välja kirjutatud retseptid või tervishoiuasutuses manustatud ravimid* | 2012-07-31 | – |
| EMCALL | Emergency call center case data<br>*Juhtumi andmed häirekeskusest* | 2012-07-31 | – |
| EMCALL_ENTER | Entered emergency call center case data<br>*Sisestatud häirekeskuse juhtumi andmed* | 2012-07-31 | – |
| EMDUTY | Emergency call-out and dispatch details<br>*Hädaabikutse ja väljasõidukorralduse ning kutse täitmise andmed* | 2012-07-31 | – |

| Section code | Section name | Valid since | Observed in dataset |
|---|---|---|---|
| EMDUTY_ENTER | Entered emergency call-out and dispatch details<br>*Sisestatud hädaabikutse ja väljasõidukorralduse ning kutse täitmise andmed* | 2012-07-31 | – |
| EMPLSCHEDULE | Ambulance crew member schedule<br>*Kiirabibrigaadi liikme graafik* | 2012-07-31 | – |
| FIRSTAID | First aid before ambulance arrival<br>*Patsiendi abistamine enne kiirabibrigaadi kohalejõudmist* | 2012-07-31 | – |
| HAZARD | Risk of infection or contamination and/or other isolation need<br>*Nakkus- ning kontaminatsioonioht ja/või muu isolatsiooni vajadus* | 2012-07-31 | – |
| INJURY | Trauma<br>*Trauma* | 2012-07-31 | – |
| OUTSOURCE | Use of additional assistance<br>*Täiendava abi kasutamine* | 2012-07-31 | – |
| PATOBJ | Patient objective status<br>*Patsiendi objektiivne staatus* | 2012-07-31 | – |
| PATOBS | Additional patient data<br>*Täiendavad patsiendi andmed* | 2012-07-31 | – |
| POISON | Poisoning<br>*Mürgistus* | 2012-07-31 | – |
| REJECT | Refusal of further assistance<br>*Patsiendi edasisest abist keeldumine* | 2012-07-31 | – |
| RELATEDDOC | Reference to another document for the same patient and case<br>*Viide teisele dokumendile, mis on sama patsiendi sama juhtumiga seotud* | 2012-07-31 | – |
| TCOTHER | Other important additional information<br>*Muu oluline lisainfo* | 2012-07-31 | – |
| TIMECRITICAL | Time-critical patient data<br>*Aeg-kriitilised andmed patsiendi kohta* | 2012-07-31 | – |
| TRANSP | Mode of transport<br>*Transpordi viis* | 2012-07-31 | – |
| VEHCLSCHEDULE | Ambulance crew and vehicle relation<br>*Kiirabibrigaadi ja liiklusvahendi seos* | 2012-07-31 | – |
| REASON | Justification<br>*Põhjendus* | 2013-02-01 | – |
| TD | Health declaration<br>*Tervisedeklaratsioon* | 2013-02-01 | – |

| Section code | Section name | Valid since | Observed in dataset |
|---|---|---|---|
| TTDEC | Health certificate decision<br>*Tervisetõendi otsus* | 2013-02-01 | – |
| TTHEALTH | Person's health data<br>*Isiku terviseandmed* | 2013-02-01 | – |
| TTPROC | Additional findings and examinations for health certificate<br>*Tervisetõendi täiendavad leiud ja uuringud* | 2013-02-01 | – |
| CLIN_DGN | Clinical diagnosis<br>*Kliiniline diagnoos* | 2014-03-05 | ✓ |
| DEC | Decision<br>*Otsus* | 2014-03-05 | ✓ |
| PAT_DGN | Pathomorphological diagnosis<br>*Patomorfoloogiline diagnoos* | 2014-03-05 | ✓ |
| PAT_PROC | Pathology examinations<br>*Patoloogia uuringud* | 2014-03-05 | ✓ |
| RG_PROC | Radiology examinations<br>*Radioloogilised uuringud* | 2014-03-05 | ✓ |
| CALLINGSCHEDULE | Ambulance call schedule<br>*Kiirabibrigaadi kutsungi graafik* | 2015-04-16 | – |
| DENTCLINOBS | Dental clinical observation<br>*Hambaravi kliiniline vaatlus* | 2015-01-01 | – |
| DENTDISE | Dental care case<br>*Hambaravi haigusjuhtum* | 2015-01-01 | – |
| HEALTHSTATE | Patient-stated health data<br>*Patsiendi ütluspõhised terviseandmed* | 2015-01-01 | – |
| RANKCHEDULE | Ambulance crew level schedule<br>*Kiirabibrigaadi taseme graafik* | 2015-04-16 | – |
| STATUSCHEDULE | Ambulance crew status schedule<br>*Kiirabibrigaadi oleku graafik* | 2015-04-16 | – |
| DIR_PURPOSE | Purpose of referral<br>*Suunamise eesmärk* | 2016-12-12 | ✓ |
| DIR_SERVICE | Referral to service<br>*Suunamine teenusele* | 2016-12-12 | ✓ |
| DRUGSCH | Medication regimen<br>*Ravimiskeem* | 2016-06-17 | ✓ |
| ENDO | Endoscopy examinations<br>*Endoskoopia uuringud* | 2016-04-01 | ✓ |
| INTOL | Known drug side effects<br>*Teadaolevad ravimite kõrvaltoimed* | 2016-06-17 | – |

| Section code | Section name | Valid since | Observed in dataset |
|---|---|---|---|
| KOLO | Colonoscopy examinations<br>*Koloskoopia uuringud* | 2016-04-01 | ✓ |
| DEATH_CAUSE | Cause of death<br>*Surma põhjus* | 2017-12-12 | – |
| ID | Identification<br>*Identifitseerimine* | 2017-12-12 | – |
| P_DEATH | Perinatal death<br>*Perinataalsurm* | 2017-12-12 | – |
| P_DEATH_CAUSE | Cause of perinatal death<br>*Perinataalsurma põhjus* | 2017-12-12 | – |
| INJ_FALL | Injury resulting from a fall<br>*Kahjustusega lõppenud kukkumine* | 2018-11-30 | – |
| INT_SUR | Planned surgery<br>*Planeeritav operatsioon* | 2018-11-30 | – |
| METRICS | Anthropometric measurements<br>*Antropomeetrilised näitajad* | 2018-11-30 | – |
| NURSE_SUM | Nursing services provided to the patient<br>*Patsiendile osutatud õendusabiteenuse tegevused* | 2018-11-30 | – |
| NURSING_CONSULT | Overview of provided consultations and home visits<br>*Ülevaade osutatud konsultatsioonidest ja koduvisiitidest* | 2018-11-30 | – |
| PIC_REFR | References to used image links<br>*Viited kasutatud pildiviitadele* | 2018-04-05 | – |
| RISK_FACTOR | Risk factors<br>*Ohutegurid* | 2018-11-30 | ✓ |
| EC | EMS cards<br>*Kiirabikaardid* | 2020-10-29 | – |
| REF | Referral letters<br>*Saatekirjad* | 2020-10-29 | – |
| BKG | Bookings<br>*Broneeringud* | 2021-02-04 | – |
| CLOSED | Closed documents<br>*Suletud dokumendid* | 2024-09-09 | – |

#### 4 Section type introduction timeline (Figure S5)

Figure S5 complements Table S1 by showing temporal clustering in the introduction of section types across the broader Estonian CDA R2 specification landscape.

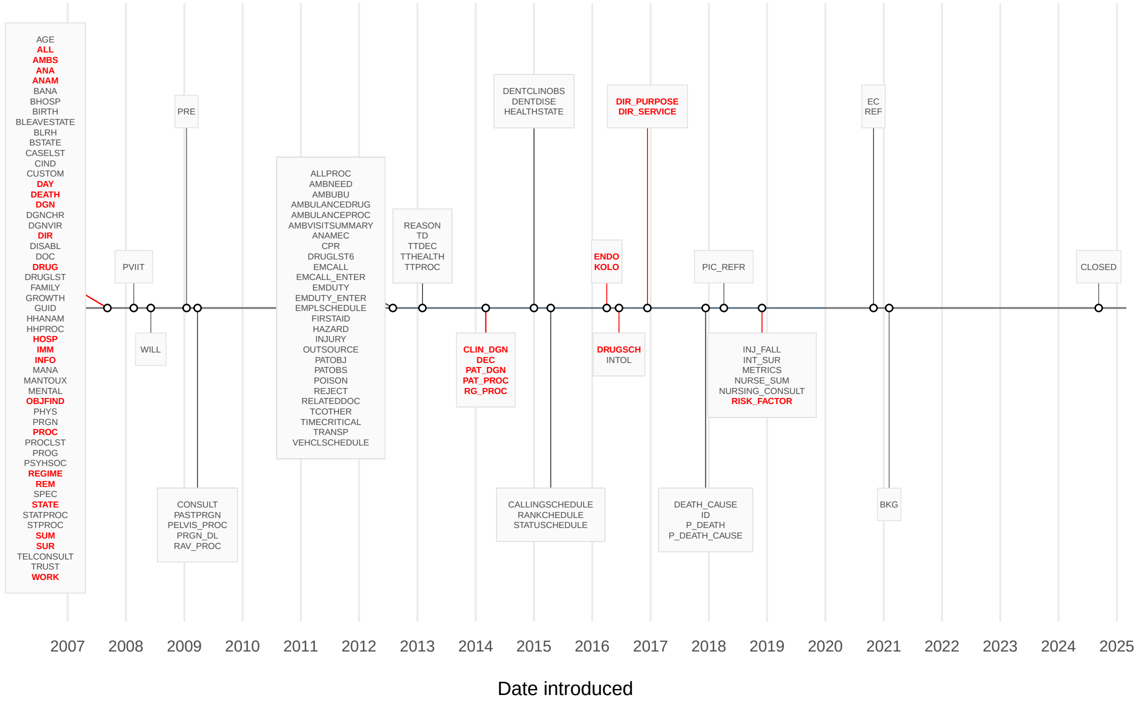


**Figure S5.** Section type introduction timeline for the broader section catalogue. Section types highlighted in red are those observed in the 2012–2019 study dataset (marked ✓ in Table S1).

### 5 XPath compaction notation for within-section heatmaps (Table S2)

To keep the XPath-level heatmaps in Figure 4 of the main article and in Figures S6–S8 legible, sibling attributes that share a parent element and whose per-section mean occurrences coincide year by year (to within the display-rounding tolerance of the printed cell value) are merged into a single "bundle" row, labelled in the `@{...}` form within the partial XPath. A bundle is formed only when at least two of the listed sibling attributes are actually present under that parent and their per-year means agree; otherwise the attributes remain as separate rows. Bundling is applied in two passes: first-level bundles are formed from co-occurring sibling attributes (Table S2); a second pass then collapses the `@{CD}` bundle together with a sibling `@type` attribute into the second-level bundle `@{CD+type}`.

The bundle labels reflect the underlying HL7 v3 / CDA datatypes and Reference Information Model (RIM) attribute groupings: `@{CD}` names the four components of the HL7 v3 Concept Descriptor (CD) datatype; `@{CC+MC}` and `@{CC+DC}` capture the two common pairings of the RIM `classCode` attribute with either `moodCode` or `determinerCode`; `@{HL7META}` groups attributes that identify the document and its underlying HL7 model rather than carrying clinical content; and `@{inclusive+value}` reflects the boundary pair used inside interval-typed elements such as `value/high` and `value/low`.

**Table S2.** XPath compaction bundles used in the within-section heatmaps (Figure 4 of the main article; Figures S6–S8 of the supplementary). The component-attributes column wraps where needed.

| Bundle label | Component attributes | Interpretation |
|---|---|---|
| `@{CD}` | `@code`, `@codeSystem`, `@codeSystemName`, `@displayName` | HL7 v3 Concept Descriptor (CD) datatype |
| `@{CD+type}` | `@{CD}` with sibling `@type` | Second-level bundle (typed coded concept) |
| `@{CC+MC}` | `@classCode`, `@moodCode` | RIM class / mood pairing |
| `@{CC+DC}` | `@classCode`, `@determinerCode` | RIM class / determiner pairing |
| `@{HL7META}` | `@HL7-ClassName`, `@HL7-Domain`, `@realmCode` | Document and HL7-model identifiers (not clinical content) |
| `@{inclusive+value}` | `@inclusive`, `@value` | Interval-boundary pair (e.g. under `value/high`) |

HL7 RIM attributes referenced above: `classCode` identifies the RIM class of the element (e.g. Act, Observation, Encounter); `moodCode` identifies its mood (e.g. event, intent, request); `determinerCode` distinguishes an instance from a kind for entity-typed classes (e.g. a specific specimen versus a specimen kind); `realmCode` identifies the geographic or regulatory realm of the document (`EE` for Estonia).

### 6 Additional XPath-level heatmaps for inpatient discharge summaries (Figures S6–S8)

Figures S6–S8 extend the main-text XPath-level analysis of the laboratory investigations (`ANA`) section to three further high-prevalence sections in inpatient discharge summaries: final clinical diagnosis (`DGN`, Figure S6), hospital stay (`HOSP`, Figure S7), and examinations and procedures (`PROC`, Figure S8). All heatmaps use the same visual encoding: rows denote compacted XPaths, columns denote years, cell labels report the mean number of occurrences per section instance, and colour encodes a contrast-enhanced symmetric log-derived transform of the same quantity centred at 1. The same XPath compaction notation is used throughout; bundle labels are defined in Section 5 (Table S2).

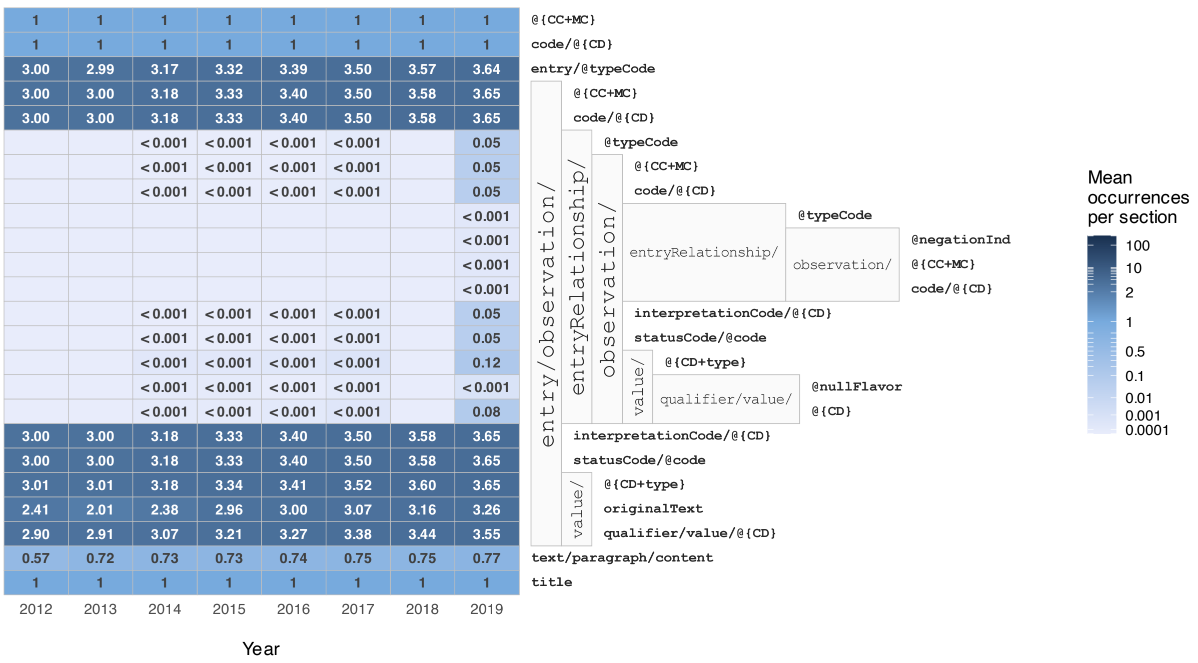


**Figure S6.** Year-specific mean XPath occurrence within the final clinical diagnosis (DGN) section of inpatient discharge summaries.

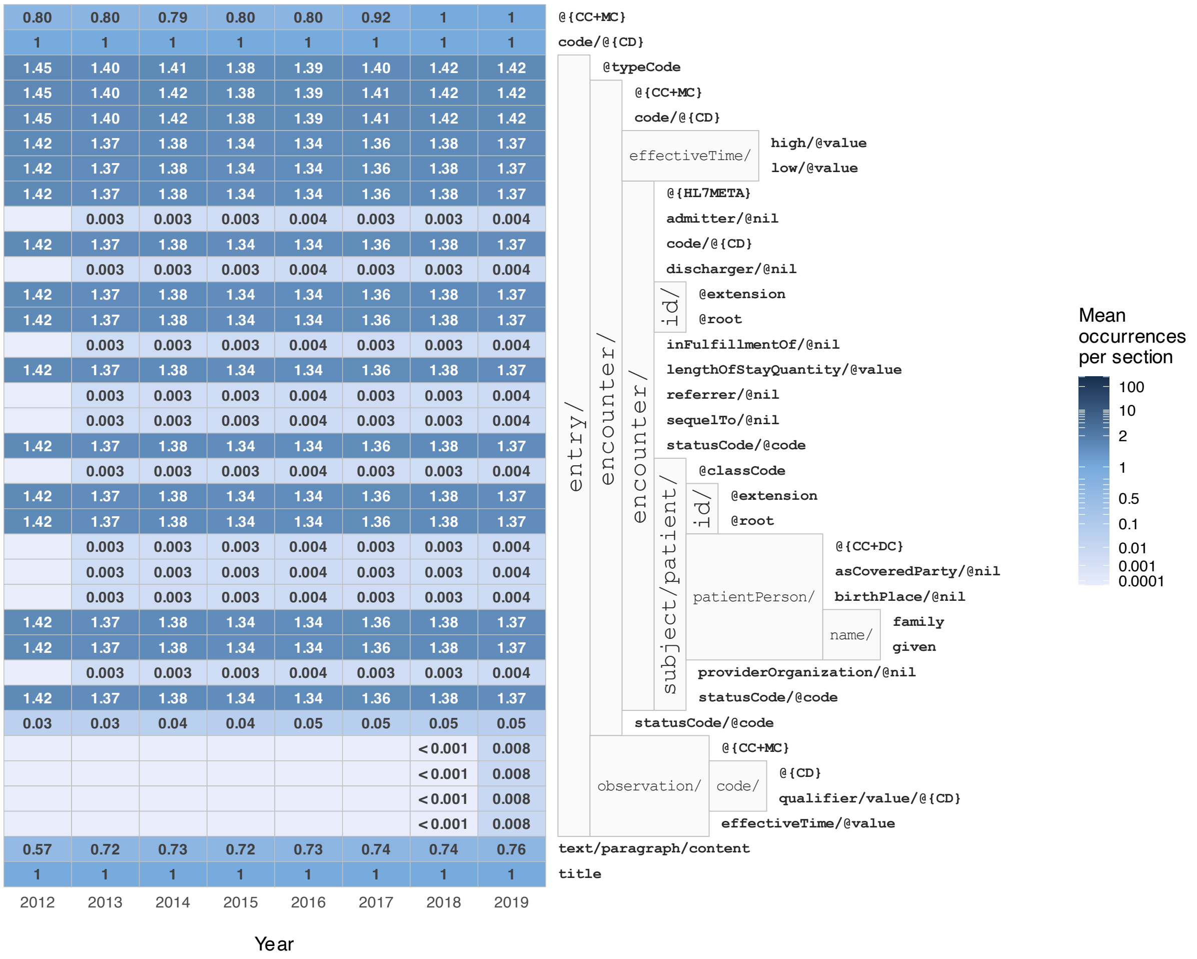


**Figure S7.** Year-specific mean XPath occurrence within the hospital stay (HOSP) section of inpatient discharge summaries.

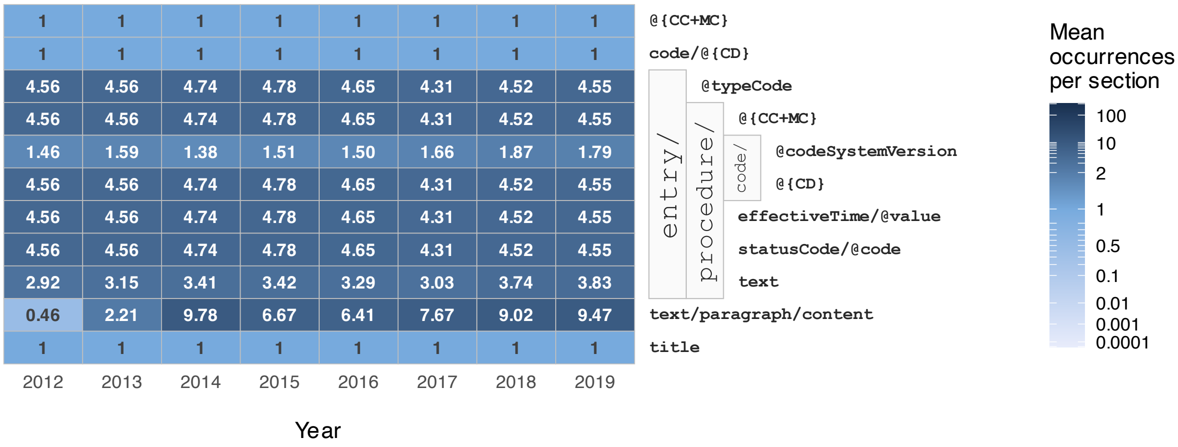


**Figure S8.** Year-specific mean XPath occurrence within the examinations and procedures (PROC) section of inpatient discharge summaries.
